# Modeling disease progression in spinocerebellar ataxias

**DOI:** 10.1101/2024.05.29.24308162

**Authors:** Elisabeth Georgii, Thomas Klockgether, Heike Jacobi, Tanja Schmitz-Hubsch, Tetsuo Ashizawa, Sheng-Han Kuo, Tim Elter, Marie Piraud, ESMI study group, EUROSCA study group, RISCA study group, CRC-SCA study group, SCA-Registry study group, Jennifer Faber

**Author notes:** Corresponding author: Dr. Jennifer Faber, Deutsches Zentrum für Neurodegenerative Erkrankungen (DZNE); Venusberg-Campus 1/99; 53127 Bonn; Germany.

## Abstract

**Background and objectives:** The most common autosomal-dominantly inherited spinocerebellar ataxias (SCA), SCA1, SCA2, SCA3 and SCA6, account for more than half of all SCA families. Disease course is characterized by progressive ataxia and additional neurological signs. Each of these SCAs is caused by a CAG repeat expansion, leading to an expanded polyglutamine stretch in the resulting type-specific protein. To comparatively investigate determinants of disease progression, we analyzed demographic and genetic data and three-year clinical time courses of neurological symptoms. The aim was to provide tailored marker candidates and prediction models to support type-specific clinical monitoring and trial design.

**Methods:** To analyze relationships among the different neurological symptoms, we examined co-occurrence patterns of deterioration events. Predicting disease progression was treated as a survival analysis problem.

**Results:** The data set contained 1538 subjects from five different longitudinal cohorts and 3802 visits. The pattern of neurological symptoms that showed progression varied with the SCA type. Mining of the progression data revealed the Scale for the Assessment and Rating of Ataxia (SARA) sum score to be the most representative descriptor of disease progression, reflecting progression of the majority of the other included symptoms. We trained models for predicting the progression of each neurological symptom for each SCA type from genetic features, age and symptoms at the baseline visit. The most universal predictors included the SARA sum score, gait and the CAG repeat length of the expanded allele. Finally, deterioration in disease staging was studied in detail: For the milestones of deterioration, (i) the need to use walking aids and (ii) the requirement to use a wheelchair, we discovered common as well as diverging predictive markers. For clinical interpretability, a decision tree was built to indicate the probability of progression within 3 years in dependence of the top predictive features.

**Discussion:** Data-driven approaches are potent tools to identify the main contributing features of progression prediction. Progression events for the disease stage were predictable from the baseline neurological status. Remarkably, a limited number of features had predictive importance, and only few were shared among all four SCA types, including gait and the SARA sum score, confirming the need for type-specific models.

## Introduction

Spinocerebellar ataxias (SCAs) are rare neurological diseases with autosomal dominant inheritance and a clinical onset in adult life. Patients suffer progressive loss of balance and coordination accompanied by slurred speech. By now, more than 50 genetically different SCAs are known, among which SCA1, SCA2, SCA3 and SCA6 belong to the most common. ^1, 2^ Each of these SCAs is caused by a translated CAG repeat expansion in the respective gene resulting in an expanded polyglutamine (PolyQ) in the protein. ^1^ Disease-modifying therapies applying gene silencing strategies are currently developed, and first human safety trials with antisense oligonucleotides have started. A comprehensive knowledge of the disease course is a core requirement for the planning and design of clinical trials. Disease course of SCAs has been studied mainly with two clinical rating assessments, the Scale for the Assessments and Rating of Ataxia (SARA), which provides a measure of ataxia severity ^3^, and the Inventory of Non-Ataxia Signs (INAS) ^4^, which assesses neurological symptoms other than ataxia, such as e.g. spasticity, rigidity or dysphagia. ^5–7^ A prediction of disease progression based on the current status could improve stratification of patients for clinical trials as well as patient counselling. The main objective of this work was to develop a model that predicts progression in SCA1, SCA2, SCA3, and SCA6 based on demographic data, CAG repeat length and baseline clinical measures. We were particularly interested in predicting risks of reaching milestones of high patient relevance. To this end, we compiled longitudinal data of more than 1500 SCA1, SCA2, SCA3 and SCA6 mutation carriers from four European and one US SCA cohorts. For the analysis, we chose a purely data-driven approach.^8^

## Methods

### SCA patient cohorts

We compiled clinical data from four European observational studies in the most common SCAs, namely EUROSCA (N=506) ^6, 9^, RISCA (N=138) ^7, 10^, ESMI (N=310) ^11^ and SCA-Registry (N=265), as well as the US cohort CRC-SCA (N=337) ^5^. All participants were assessed using a cohort-specific, but largely overlapping standardized protocol that included demographic data and genetic characteristics, such as age and CAG repeat length. Clinical assessments included SARA ^3^ (to grade ataxia severity) and INAS ^4^ (to indicate non-ataxia symptoms). SARA consists of eight rated items, namely gait (0-8), stance (0-6), sitting (0-4), speech (0-6), finger chase (0-4), nose-finger test (0-4), fast alternating hand movements (0-4) and heel-shin slide (0-4), yielding a sum score ranging from 0 (absence of ataxia) to 40 (most severe ataxia). INAS assesses the presence vs. absence of the following neurological signs: hyperreflexia, areflexia, extensor plantar reflex, spasticity, paresis, muscle atrophy, fasciculations, myoclonus, rigidity, chorea/dyskinesia, dystonia, resting tremor, sensory symptoms, urinary dysfunction, cognitive dysfunction and brainstem oculomotor signs, the latter comprising ophthalmoparesis on horizontal and/or vertical gaze and/or slowing of saccades. The INAS count gives the number of neurological signs as a global measure of non-ataxia involvement, ranging from 0 to 16. In addition, the following INAS items were assessed: downbeat nystagmus on fixation, gaze evoked-nystagmus horizontal and vertical testing, square wave jerks on fixation, hypo– and hypermetric saccades, broken up smooth pursuit, visual acuity, diplopia as well as dysphagia (the latter two with ranges 0 to 3, all other 0/1). ^4^ Disease stages were defined following Klockgether et al. ^12^ as 0 = no gait difficulties, 1 = ataxic, defined by the presence of gait difficulties, 2 = loss of independent gait with the need to use walking aids, 3 = confined to wheelchair. The US cohort used a more fine-grained staging system, that was mapped onto the stages 0 – 3 as done previously ^13^. A comprehensive list of items as well as an overview of the mapping can be found in the Supplementary data (Table S2). We performed a Kruskal-Wallis test with Dunn’s posthoc test, p<0.01, to compare age and disease characteristics between SCA1, SCA2, SCA3 and SCA6. Written informed consent in accordance with the declaration of Helsinki was obtained from all participants.

### Analyses

We excluded two subjects with invalid genetic information and 16 subjects with invalid entries in neurological scales. The data were brought into a common format, and the disease stage annotation was harmonized (Table S1). According to the objectives of this work, we grouped the data into progression data (scores at up to five follow-up visits within 3 years) and input data (possible predictor variables, including SARA and INAS assessments at baseline, genetic features and age). Only observed values of progression data were used to label patients and to train and evaluate prediction models. For each SARA or INAS item, an increase in the assigned value observed during any follow-up visit compared to the baseline visit corresponds to a deterioration of the symptom and indicates a progression of the disease. To analyze relationships among the different neurological symptoms, we examined co-occurrence patterns of deterioration events. The task of predicting disease progression for a specific neurological symptom was treated as a survival analysis problem. We consider progression-free survival, which is equivalent to maintaining a certain capability without deterioration in the assessed scale. For that, we used fixed time windows of 0.5, 1, 1.5, 2 and 3 years after the baseline visit, ignoring minor time shifts of up to two months in the dates of visit appointments. The information on events and times was set into relation with patient characteristics at the baseline visit by applying survival forests, which were selected as the best performing approach among four survival analysis methods applied to each of the individual SCA types. The performance was evaluated by the concordance index on the same test sets in four-fold cross-validation. The transitions between the most relevant clinical milestones, the deterioration of disease stages (namely loss of free walking ability/need to use walking aids and loss of ambulation/need to use a wheelchair), were modeled separately to specifically identify features guiding each transition. For each training set, a bootstrapping approach was used to select the most predictive features on hundred resampled instances of the training set. All analyses were performed using R, version 4.1.0. For detailed information on data handling and visualization, association rule mining, progression prediction and feature selection as well as the respective R packages see Supplement (6.1).

## Results

### Clinical and demographic characteristics

The combined SCA cohorts comprised 1538 subjects, including 147 healthy controls, with a total of 3802 visits. There were maximally six visits per subject, within a time frame of maximally three years. Clinical and demographic characteristics are summarized in Table 1.

**Table 1:** Summary characteristics for SCA1, SCA2, SCA3 and SCA6 mutation carriers as well as healthy controls from the collected data.

| Type | Subjects<br>(#) | Visits<br>(#) | Cohorts<br>(#) | Age<br>(median) | Male<br>(%) | SARA<br>at baseline<br>(median) | Maximum annual<br>SARA progression<br>(median) | CAG repeats<br>expanded allele<br>(median) | CAG repeats<br>normal allele<br>(median) |
| --- | --- | --- | --- | --- | --- | --- | --- | --- | --- |
| SCA1 | 276 | 680 | 4 | 44 | 50 | 10 | 1.86 | 47 | 30 |
| SCA2 | 306 | 850 | 4 | 45 | 49 | 13 | 1.58 | 39 | 22 |
| SCA3 | 589 | 1467 | 4 | 48 | 49 | 10 | 1.50 | 70 | 22 |
| SCA6 | 220 | 606 | 4 | 64 | 50 | 13 | 1.02 | 22 | 12 |
| Controls | 147 | 199 | 1 | 44 | 50 | 0 | 0.00 | - | - |

### SCA population shows a continuous spectrum of disease severity

Based on their clinical time courses, the subjects in the compiled data collection form a continuum from healthy state to non-severe disease to severe disease stages (Figure 1). Patients with a transition from a non-severe (disease stages 0-1) to a severe state (disease stages 2-3) during the observed time period are located at the interface between both states (Figure 1 A). The only visible structure among the subjects are three different stripe-like clusters, each one separately replicating the common disease stage gradient. Those clusters are directly related to the number of missing values in the original data and the different composition of the RISCA cohort with a focus on pre-ataxic mutation carriers (Figure S1 B-C). (Figure S1 D). Apart from that, the cohorts mix and do not show any batch effects. Similarly, the different SCA types are well distributed across clusters and disease severity levels, indicating that overall disease severity and data acquisition standards do not differ between SCA types (Figure S1 E).

**Figure 1:**
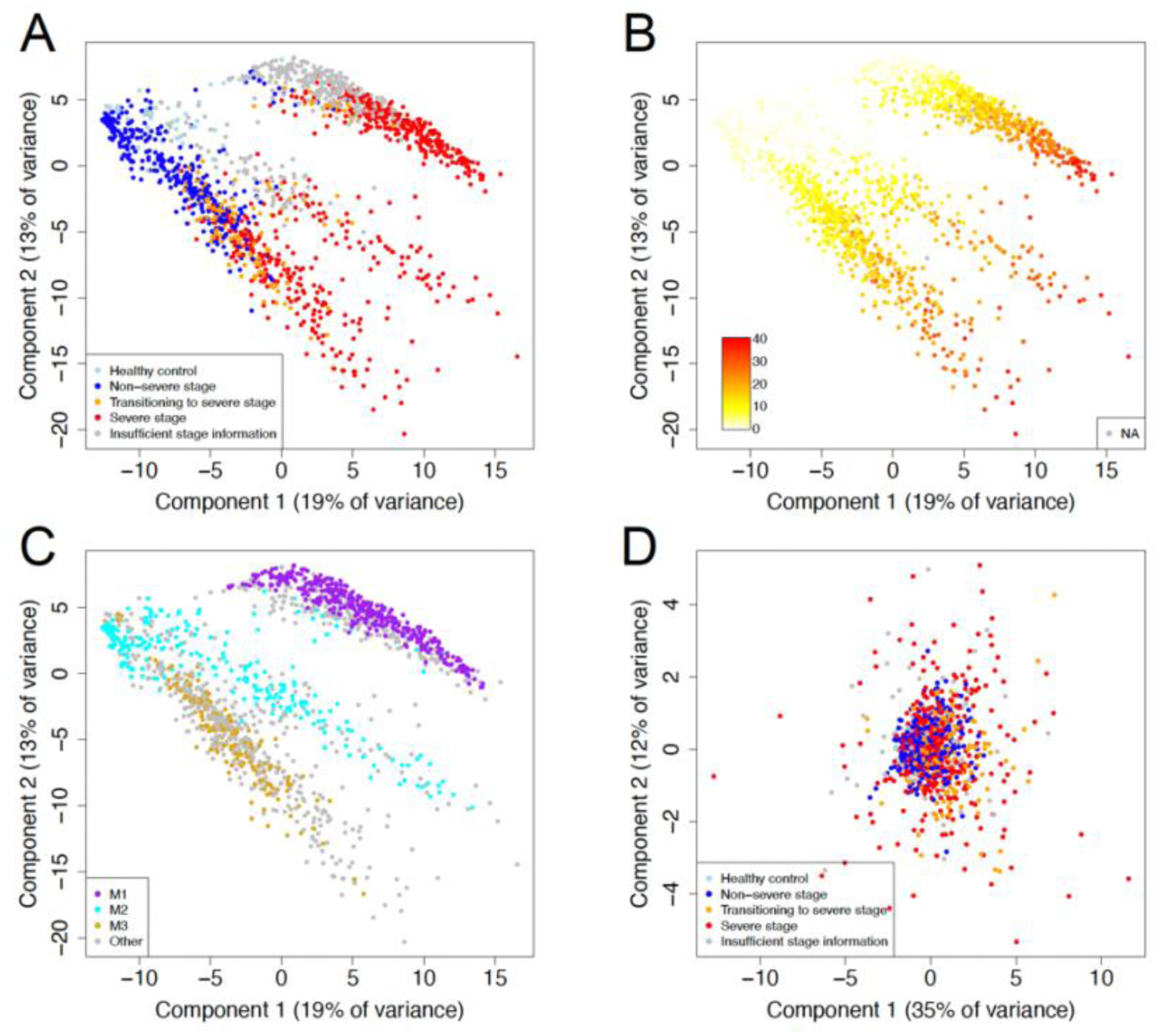
Overall visualization of subjects based on principal component analysis of clinical time courses across all neurological scales. A. Coloring by disease stage annotation (Healthy: control subject; Non-severe stage: stage 0 or 1, corresponding to normal status or ataxic gait, but ability of free walking, at baseline visit and all follow-up visits; Severe stage: stage 2 or 3, corresponding to the need to use walking aids or wheelchair, already at baseline visit; Transitioning to severe stage: stage 0 or 1 at baseline visit and stage 2 or 3 at a follow-up visit within three years from the baseline visit; Insufficient stage information: SCA subject with non-severe stage at baseline and at follow-up visits up to one year, but with no further follow-up visits afterwards). B. Coloring by SARA sum score at the baseline visit. Both coloring schemes show a gradient from the top left to the bottom right. C. Coloring by the major missing value bins related to the number of missing values (see Figure S1B). The visible stripe-like clusters (A-C) are explained by the number of missing values in the input data (C). D. Principal component visualization of subjects based on maximum annual progression rates of the SARA sum score and individual SARA items. Coloring by disease stage annotation like in A. Here, no clusters can be identified.

The disease stage gradient aligns with the SARA sum score at the baseline clinical visit, (Figure 1 B) but not with the SARA sum score annual progression (Figure S1 F). This means, the baseline score is indicative of disease stage, but the rate of SARA progression does not differ between different stages of the disease (Figure 1 D, Figure S2). Even the subjects that transitioned to the severe stage within the three years do not appear as a separate cluster (Figure S2).

### SARA sum score is the most representative descriptor of disease progression

Although the SARA progression rates did not reveal distinct patient groups, the SARA sum score progression was most representative of the overall disease progression. Among the neurological scales (including SARA items, SARA sum score, INAS items, INAS count and disease staging), a deterioration of any single symptom likely implied a deterioration of the SARA sum score. These relationships, called association rules (Section 6.1), were found with high confidence scores across patients from all SCA types. For disease staging and the individual SARA items, the rule confidence scores always lay above 92 %. For instance, in 536 of the 564 cases, where the SARA gait score worsened, also the SARA sum score worsened, yielding a confidence of 95 % for the rule. Also, a deterioration of individual INAS items was always reflected in a deterioration of the SARA sum score in at least 80 % of the cases. Among the INAS items, tremor and muscle atrophy implied a SARA sum score deterioration with the largest confidence (94 %). In contrast, the INAS count score reflected only deterioration of fifteen single INAS items with confidence scores above 80 % in the data. This means, although the SARA sum score aggregates only SARA items, it is a better descriptor of the overall disease state with respect to INAS symptoms than the INAS count. Many combinations of items were perfectly associated (i.e., with a confidence score of 100 %) with the SARA sum score, the INAS count or single items. For instance, if SARA item *heel-shin slide* and INAS item *broken up smooth pursuit* deteriorated, the SARA sum score deteriorated.

### Future disease progression events are predictable from the current neurological status

#### Progression frequency

An overview of the relative progression frequencies within 3 years of all considered neurological items is given in Figure S4. In general, SCA6 differed from the other SCA types, exhibiting lower deterioration frequencies for the majority of INAS items. Overall, the individual SARA items, which include severity ratings, naturally showed a much larger deterioration frequency in the observed 3-year period than the INAS items coded as absent or present. Only the INAS count reflected deterioration of a larger fraction of patients.

#### Progression predictability

We compared four modeling approaches for the prediction of transition to higher disease stage. Among these, the survival forest approach was the only one that performed consistently well for all the SCA types (Figure S5). Consequently, this approach was chosen for all following analyses. The performance of deterioration predictions varied with SCA type and neurological item, but was above random for almost all items in each SCA type (Table S1, Figure S6). With the largest number of recorded patients, SCA3 showed the best overall prediction performances. Disease stage predictability on all cross-validation test sets was between 0.62 and 0.95 for all SCAs.

#### Identification of the top predictive features

Only a small number of features had a high importance for the prediction of disease stage progression. The baseline disease stage, the baseline SARA sum score and the baseline SARA gait score were among the top five predictors in each SCA type (Figure 2). Furthermore, SARA stance showed increased relevance for SCA1 and SCA2, whereas SCA6 got the largest importance score for INAS dysphagia among the SCA types, and the lowest importance score for the number of repeats in the expanded allele. A comprehensive overview for each SCA type is given in Figure S7). We used Sankey diagrams to visualize the most predictive items for the progression of each neurological item (Figure S8). For each item, the three top predictors are given. The majority of scales and items had their own baseline value among the top three predictors. Beyond those evident predictors, there were many cross-relationships among items. Overall, the baseline score of SARA gait was among the most universal predictors for SCA2, SCA3 and SCA6 (Figure S8, Table S1). Likewise, the number of repeats in the expanded allele was a main predictor for SCA1 and SCA3.

**Figure 2:**
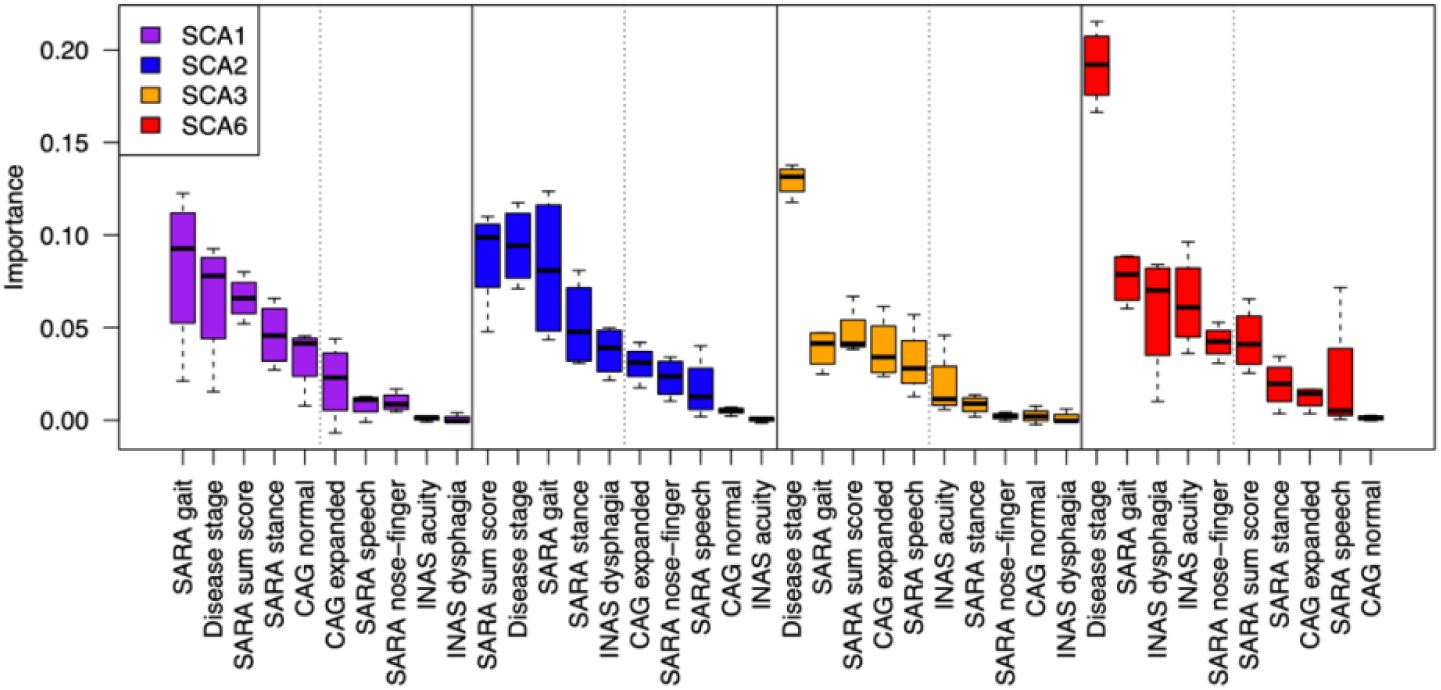
Top important input features for predicting disease stage progression in SCA1, SCA2, SCA3 and SCA6. The deterioration of disease stage (normal, gait disturbances, walking aids, wheel chair) marks key clinical milestones. Neurological measurements at baseline visit, age and genetic information were the input features for the prediction of disease stage progression. For every input feature, the importance values for the prediction of disease stage deterioration were assessed in each SCA separately and for each SCA type, the features with the top five median importance values were selected. For cross-type comparisons the top features of all SCA types were combined and sorted from high to low according to their importance within each SCA type, respectively. The top five features for the respective SCA type are to the left of the dotted line. This is a condensed representation of the importance plots across all input features (Figure S7). Boxplots indicate the distribution of feature importance values assigned by survival forests in four-fold cross-validation (bold black line: median, box: 0.25 and 0.75 quantile, whiskers: minimum and maximum value). CAG expanded: number of CAG repeats in the expanded allele, CAG normal: number of CAG repeats in the normal allele, a comprehensive overview of single INAS and SARA items is given in Table S2.

#### Each disease stage transition is characterized by specific clinical marker*s*

In the disease course of SCAs, two clinical milestones of high impact are the loss of the ability of free walking, coming with the need to use walking aids (transition from stage 1 to stage 2), and the need to use a wheelchair (transition from stage 2 to stage 3). Within our data set, the largest number of such observed transitions was available for SCA3 (N = 57 and N = 39, respectively; Figure S4, Table S4), so we describe these results in more detail in the following paragraphs.

#### Transition from disease stage 1 to disease stage 2 in SCA3: Loss of free walking ability

The loss of free walking ability could be predicted for SCA3 from all the baseline features with a median test performance of 0.79. In spite of the reduced number of training examples for the specific transition, this test performance is similar to the test performance for the overall stage progression including all disease stage transitions (Table S1). To highlight the most central predictive markers, we determined features that were robustly identified to be among the top five predictive in multiple training sets during cross-validation (6.1). Age, INAS count, presence of fasciculations, CAG repeats of the normal allele, SARA items finger chase, gait and stance as well as SARA sum score were the main features that contributed to predicting progression, i.e. loss of free walking, within three years (represented by a column on the left-hand side in Figure 4 A, a color key indicating the range of values is given below each column, respectively). The heatmaps allow interpretations in horizontal as well as vertical directions. Following a horizontal line, one particular combination of features can be related to the respective predicted 3-year outcome on the right-hand side, represented as the probability to keep the ability of free walking within three years. Advanced baseline scores for SARA sum, SARA stance, SARA gait and SARA finger chase corresponded to a high risk of stage transition, whereas low scores of these features corresponded to a low risk of stage transition (Figure 4 A). Furthermore, the low-risk groups were characterized by a low age at the baseline visit, a low INAS count and absence of the INAS fasciculations symptom. A vertical reading of the columns allows to identify relationships between single features and the risk of losing the ability of free walking. In general higher values in the SARA sum score as well as the SARA items gait, stance and finger chase lead to a high risk of deterioration. Here, higher values always occur in the upper portion of the respective column, which is related to a high risk of progression, as indicated by the bottom-top triangle on the right hand side of the figure. In contrast, the values of age or INAS count are mixed across the vertical of their columns, with no consistent discernible pattern. Thus, even though, they were identified as the top predictive features, the overall risk of progression depends substantially on the constellation of the other features. To further facilitate the interpretation how these predictive features in combination affect the chance of not progressing to a worse stage within three years, a decision tree was built (Figure 4 B). It shows that a SARA gait score of 2 or less and a SARA sum score of 5 or less led to the greatest probability of keeping the free walking ability, whereas a SARA gait score of at least 3 in combination with INAS fasciculations and a SARA sum score of at least 12.5 led to the lowest probability of keeping the free walking ability.

**Figure 3:**
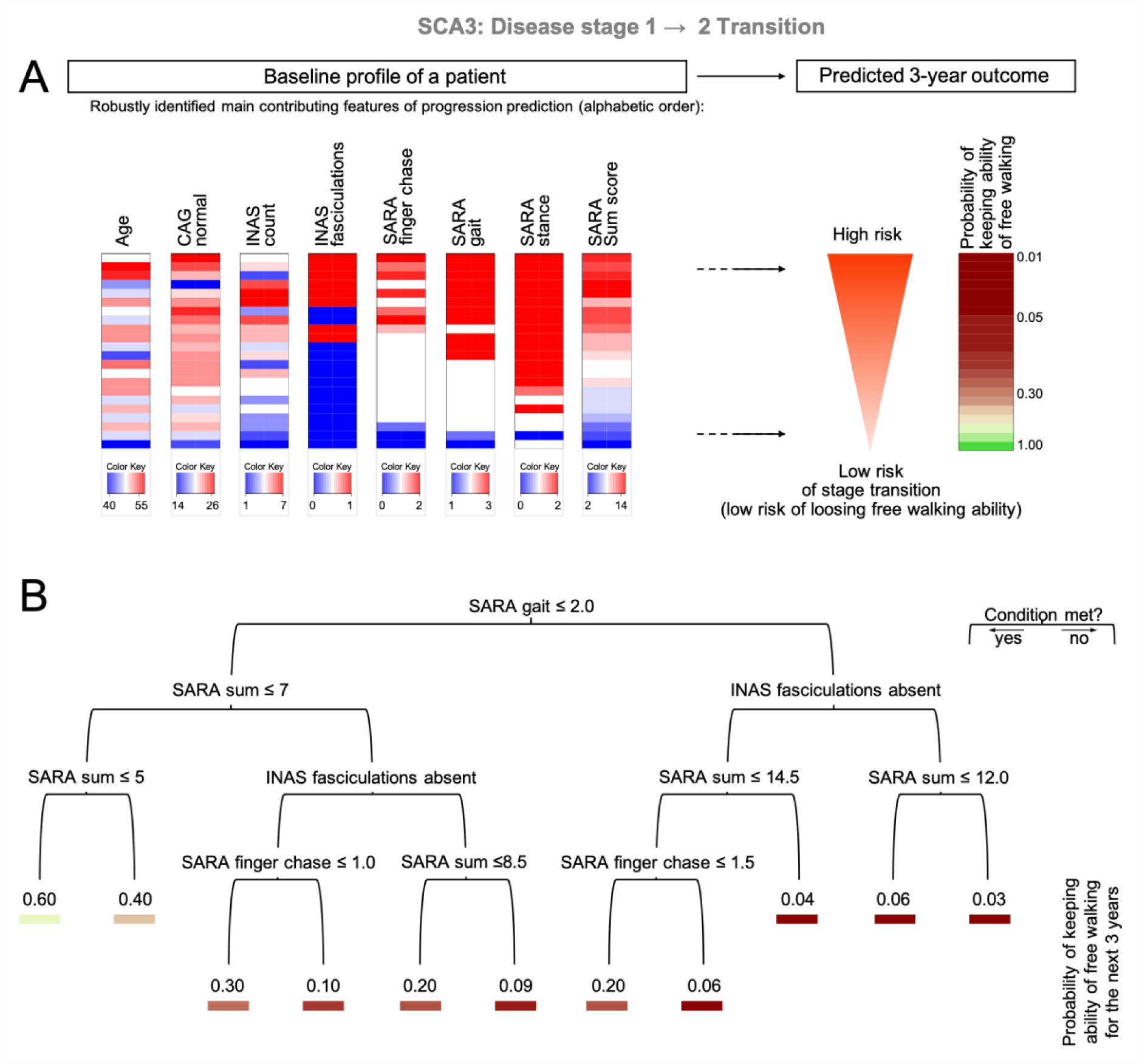
Assessing the total risk for SCA3 patients of transitioning within three years from disease stage 1 to disease stage 2 (loss of free walking ability). A. The top predictive features were selected by bootstrapping within a four-fold cross-validation (for details see Methods, Figure S9). Each column on the left hand side represents one of the features, that were robustly identified as a main contributor to the prediction for the loss of free walking ability within the next 3 years. Color coding of each feature range is indicated by the Color Key under the respective column with blue coloring indicating low values and red coloring indicating high values. The graph allows interpretations in horizontal as well as subsequently vertical reading. Following a horizontal line, one particular combination of features can be related to the respective predicted 3-year outcome by following a imagined horizontal line to the resulting probability to keep the ability of free walking within three years on the right-hand side. A vertical reading of the columns allows to identify patterns within single features related to the risk of losing the ability of free walking. It becomes obvious that higher values in the SARA items gait, stance, finger chase and SARA sum score as well as the presence of fasciculations are associated with a relatively high risk for the loss of the ability to walk freely. Here, higher values always occur in the upper portion of the respective column, which is related to a high risk (as indicated on the right-hand side). In contrast, the values of age, INAS count as well as CAG repeats of the normal allele, are mixed across the vertical of the columns, respectively, with no consistent discernible pattern. Even though they were identified as the top predictive features, the overall risk of progression depends substantially on the constellation of the other features. B. A decision tree was built from the robustly identified top predictive features to illustrate their combined effects on the probability to keep the ability of free walking within the next 3 years, derived from the mean cumulative hazard in each leaf. It is solely shown for ease of interpretation and does not constitute the full prediction model. At each inner node of the tree, the left branch satisfies the condition written at the top of the node, whereas the right branch does not satisfy it. The probability to remain in disease stage 1, or in other words keeping the ability for free walking is given at each branching end and is color coded according to the probability of keeping the free walking ability on the right-hand side in (A).

**Figure 4:**
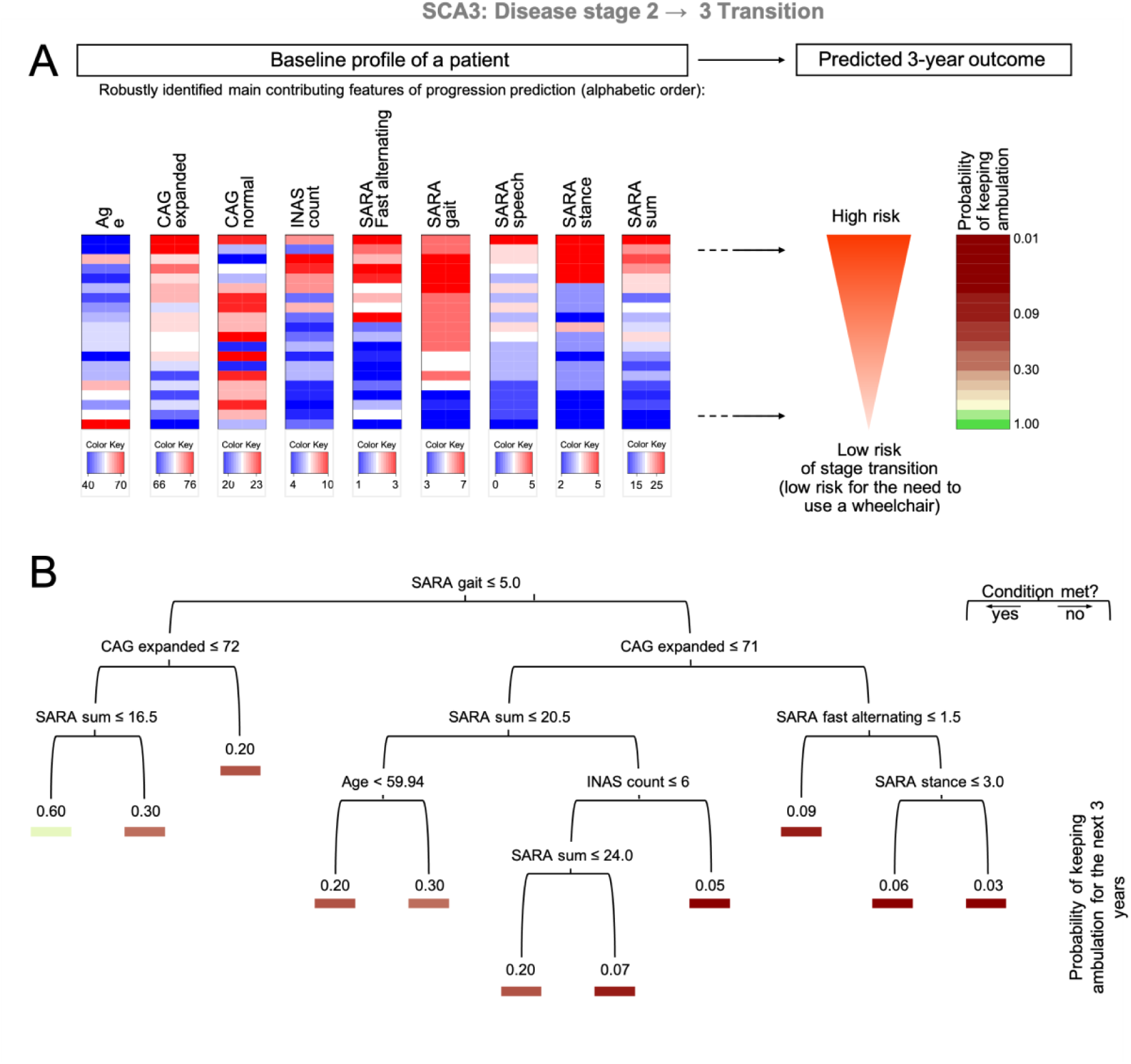
Assessing the total risk for SCA3 patients of transitioning within three years from disease stage 2 to disease stage 3 (need of using a wheelchair). A. The top predictive features were selected by bootstrapping within a four-fold cross-validation (for details see Methods, Figure S9). Each column on the left hand side represents one of the features, that were robustly identified as a main contributor to the prediction for the loss of ambulation, i.e. the need to use a wheelchair, within the next 3 years. B. Decision tree built from the robustly identified top predictive features to illustrate their combined effects on progression risk (see Figure 3 for further details on the charts).

#### Transition from disease stage 2 to disease stage 3 in SCA3: Loss of ambulation

For the transition to the need of using a wheelchair, the median cross-validation prediction performance on SCA3 patients was 0.77, which is again similar to prediction performance for the overall stage progression (Table S1). Six of the robustly identified predictive features in SCA3 patients were the same as for the loss of the free walking ability: SARA sum, SARA stance, SARA gait, the number of CAG repeats in the normal allele and age (Figure 4 A). The additionally identified predictive SARA features were SARA speech and SARA fast alternating hand movements. For both of them, low scores were associated with low risk of transitioning. Furthermore, the number of repeats in the expanded allele now also showed a clear correlation with the transitioning risk. Only in combination with high repeat numbers of the expanded allele, low age led to a high risk of needing a wheelchair (Figure 4 B). As before, SARA gait guided the basic decision in the tree, but then the number of repeats in the expanded allele played the most prominent role in this disease stage transition. High values for these two features together with large baseline scores for SARA fast alternating hand movements and SARA stance were most indicative for the highest risk of needing a wheelchair.

#### Comparison of SCA types and cross-validation of models

After focusing on stage transitions for SCA3, we now compare them to the respective stage transitions of the less common SCA types. While for SCA3 SARA item gait was the basic branching point in the decision tree for both transitions, it was SARA sum for SCA1, 2 and 6 in the transition from stage 1 to 2 (loss of free walking ability). For both SCA1 and SCA2, SARA stance got the largest importance weight after the predictors shared by all SCA types, stage, SARA sum and SARA gait (Figure 2), and had a clearer effect on the wheelchair transition (Figure S14, Figure S15) including decision trees) than on the walking aid transition (Figure S10, Figure S11). For more details, see Supplement. To further study whether a pre-selection of input features has an impact on the predictability, we compared the performance of survival forest models using all input features to models using only a subset of features. In general, regardless of the specific stage transition of interest, it was beneficial to consider the whole feature profiles during predictions (Figure S9). However, in the case of very small training datasets, such large models may overfit and feature selection may improve the prediction performance (Figure S9).

## Discussion

We analyzed a large data set of five European and US longitudinal observational studies of the most common spinocerebellar ataxias, SCA1, SCA2, SCA3 and SCA6. Using a purely data-driven approach, we found (1) that the SCA population studied represented a continuous spectrum of disease severity without evidence for separate clusters of deviating disease severity or progression rates, (2) that the SARA sum score was the most representative descriptor of disease progression, and (3) the progression to advanced disability stages can be predicted from neurological status at baseline visit, CAG repeat length and age. While previous studies that looked for predictors of disease progression concentrated on the effect of a small number of demographic and genetic factors, such as age, sex, and CAG repeat length ^6, 14^, we applied machine learning approaches that also took into account a wide spectrum of baseline clinical features related to both ataxia and non-ataxia signs and considered progression of single features as well as disease stages indicating milestones of gait deterioration.

Our finding that the studied population showed a continuous spectrum of disease severity across the four SCA subtypes was unexpected, as previous studies have emphasized differences of the clinical phenotypes and rates of disease progression between SCA subtypes.^5, 9, 15^ A plausible explanation comes from the related finding that the SARA sum score was among the top predictors of the overall disease progression. This indicates that ataxia is the major denominator of overall disease progression. Non-ataxia signs, which are differentially distributed among SCA1, SCA2, SCA3, and SCA6, do not have a sufficiently large effect on disease progression that their presence or absence would define separate clusters of deviating disease severity or progression. Rather, SARA reflected progression of the majority of non-ataxia signs included in INAS and was a better descriptor of the progression of these signs than the INAS count itself. This is in line with our previous finding derived from an analysis of the disease stage progression in RISCA and EUROSCA that SARA scores steadily worsened with increasing disease stage in all SCA subtypes.^6, 7^ Previous investigations have pointed to a number of metric weaknesses in SARA using conventional statistical methods in ataxia cohorts that were smaller and more heterogeneous than the combined cohort investigated here.^16, 17^ Our results underscore the usefulness of SARA to assess overall progression across disease stages. Since diseases stages are defined by walking ability, it is not surprising that the SARA items related to gait and stance, as well as the SARA sum score, to which these two items strongly contribute with 14 of 40 maximal score points, were identified as important predictors. However, we also identified two non-ataxia signs, dysphagia and visual acuity, as strong predictors of disease progression. Dysphagia, which is encountered in all SCA subtypes ^18^, is a result of cerebellar dysfunction, but is particularly pronounced in patients with bulbar involvement. Thus, dysphagia is a clinical indicator of a pathologically extended disease state. In addition, dysphagia may result in malnutrition and aspiration and thereby accelerate disease progression. Indeed, a survival analysis of the EUROSCA study revealed that dysphagia was a strong predictor of shorter survival.^19^ The role of impaired visual acuity in disease progression in particular in SCA6 is new, but understandable, as poor vision impairs postural stability.^20^ This finding underscores the need to monitor vision in SCA patients and to correct impairments, if possible.

When focusing on progression prediction at item level, certain observations were to be expected, such as that the main predictors for a progression of the SARA item gait were its own baseline value, SARA sum score and CAG repeats of the expanded allele. However, in other cases, combinations emerged that were not anticipated. For example, the top three predictors for the SARA sum score in SCA3 and SCA6 were non-SARA items. With regard to disease stage deterioration in SCA3, the CAG repeat length of the normal allele was a robustly identified main contributing feature. This underlines the advantage of purely data-driven approaches, which allow to identify such unexpected patterns.

Furthermore, we highlighted specific prediction models for two transitions of high clinical impact in the disease course: the loss of free walking ability and the loss of ambulation. Even though the overall number of subjects in this pooled analysis was impressively high, numbers of observed stage transitions were limited. Consequently, the drawn conclusions need to be interpreted with caution. As expected, higher scores of the SARA items gait and stance as well as SARA sum score were associated with a high risk to lose the ability of free walking or to lose ambulation within 3 years. However, for the loss of free walking ability, dysmetria of the upper limbs and the presence of fasciculations were associated with a higher risk, whereas for the loss of ambulation poor performance in fast alternating hand movements and dysarthria were predictive. Importantly, the risk prediction depends on the constellation of all features at once. Thus, some of the main predictive features stratify the risk for specific subgroups rather than exhibiting a global correlation. For example, only in the subgroup without fasciculations the number of CAG repeats correlated with the risk to lose free walking.

Although we were only able to highlight some examples from the wealth of results provided by machine learning, our study shows that such a data-driven analysis is feasible and represents significant added value due to the assumption-free approach. Our results demonstrate that the risk of disease progression can be predicted to some extent and that inclusion of as many disease descriptors as possible improves the prediction. Our automated feature importance analysis confirmed known medical relationships and generated new hypotheses. The combination of different cohorts was possible and proved to be useful. In particular, risk predictions became more reliable when a larger number of subjects with observed disease progression was available (see the performances in SCA3 vs. the other subtypes). Thus, it would be rewarding to enlarge cohorts with longitudinal data acquisitions and specifically observations of disease stage transitions. This would allow to evaluate and refine the models and to apply a wider spectrum of methods. Moreover, future prediction models enriched by additional biomarker data, such as fluid and imaging biomarker, might lead to decisive improvements. Such data science approaches to large-scale data have the potential to provide valuable insights for patient counseling as well as patient stratification in clinical trials and their design.

## Data Availability

All data produced in the present study are available upon reasonable request to the authors.

## Acknowledgement

We thank Prof. Matthias Schmid for the valuable discussions.

## Supplementary Data

### Supplementary Data: Methods

#### Data preprocessing and handling of missing values

We excluded two subjects with invalid genetic information and 16 subjects with invalid entries in neurological scales. The data were brought into a common format, and the disease stage annotation was harmonized (Table S3). According to the objectives of this work, we grouped the data into progression data (scores at follow-up visits after 0.5, 1, 1.5, 2 or 3 years) and input data (the possible predictor variables, including the scores at the baseline visit, genetic data and age). The input data contain 8.3 % missing values; 784 subjects (51.0 %) had no missing value in their input profile (Figure S1 A). For the predictive analyses, missing values in the input data were imputed by applying the k-nearest neighbor method kNN from the R package VIM (version 6.1.1) on the respective training data set of a cross-validation approach, using the default setting with k=5 and numFun=median. Missing values on the input data of test instances were imputed accordingly, using the median of the five nearest neighbors from the training set based on input features only. The progression data contain 75.0 % missing values. For 412 subjects (27 %), no follow-up assessments were available (Figure S1 B). All subjects with available follow-up assessments (N=1126) had missing values in specific assessments at one or several follow-up visits. However, 358 subjects (23.3 %) had all SARA assessments (sum score and individual items) for the follow-up visit after one, two, and three years after the baseline visit. Only observed values of progression data were used to label patients and to train and evaluate prediction models.

#### Definition of progression

For each neurological item, an increase in the assigned value observed during follow-up visits compared to the baseline visit corresponds to a deterioration of the symptom and indicates a progression of the disease. For each subject, we considered for prediction approaches the occurrence of progression and, if applicable, the first progression event. For dataset statistics and visualization, we also considered the maximum annual progression rate per subject. The annual progression rate with respect to a particular neurological item is defined as the score increase within one year. It can be estimated from the score differences of the follow-up visits to the baseline visit, relative to the respective time intervals. Due to the non-linear nature of most time courses, frequently showing a sigmoidal profile, taking the maximum rate estimate was more appropriate than taking the mean rate estimate.

#### Data visualization

To check the consistency among the five pooled cohorts and the potential existence of patient subgroups with deviating disease courses, the subjects of the combined dataset were visualized based on their time courses of neurological assessments. Due to the large fraction of missing values in the progression data, we worked with two representations of per-subject profiles to avoid biased conclusions: a) the maximum annual progression rate computed from observed data, b) imputed time course data for all neurological scales. For each data representation, principal component analysis (PCA) was performed using the prcomp function in R version 4.1.0. Subjects were colored according to different attributes, including SCA type, data source (cohort), disease stage transition, SARA sum score at the baseline visit and annual progression rate of the SARA sum score. In addition to PCA visualizations of the pooled data set, specific PCA visualizations were done for each SCA type. Beside PCA, the uniform manifold approximation and projection approach and multi-dimensional scaling were used as alternative visualization methods with several different distance measures also on non-imputed data, applying the umap R packages and cmdscale function in R, respectively. As they did not reveal more aspects of the progression data structure, we present only the PCA results.

#### Association rule mining

To analyze relationships among the different neurological scales, we examined co-occurrence patterns of deterioration events. Mining of coupled deterioration events for multiple neurological items can be done at different levels of granularity. The presented results focus on deterioration events at the subject level, pooling all events within the follow-up time of three years. The analysis was done with the R package arules, version 1.7-3, setting the minimum frequency to 0.01 and the target to “rules”. The resulting rules state associations between deterioration events in the antecedent (an if statement corresponding to the left-hand side of the rule) and deterioration events in the consequent (a then statement corresponding to the right-hand side of the rule). An example rule would be the following: If items A and B deteriorate, then item C also deteriorates. The applicability of such a rule is described by two measures, the support and the confidence. The support is the relative frequency of subjects that had deterioration events for the whole set of items included in the rule (in the example \{A,B,C\}). The confidence indicates which fraction of subjects that satisfy the antecedent also satisfies the consequent. In the example, it is equivalent to support(\{A,B,C\})/support(\{A,B\}). The confidence is 100 % if the rule holds without exception in the data. In practice, perfect rules that are only supported by single or very few subjects are not interesting. Thus, the search is guided by a minimum support threshold. Time-resolved associations with sets of neurological items that changed together at a specific time point can be computed in the same way by restricting the dataset accordingly.

#### Progression prediction

The task of predicting disease progression for a specific neurological symptom was treated as a survival analysis problem. In this context, we consider progression-free survival, which is equivalent to maintaining a certain capability without deterioration in the assessed scale. For each subject, we indicated whether a deterioration event occurred within the three years of observation. If that was the case, the time from the baseline visit to the first deterioration event was recorded. Otherwise, the time until the last follow-up visit was reported. For that, we used fixed time windows of 0.5, 1, 1.5, 2 and 3 years after the baseline visit, ignoring minor time shifts in the dates of visit appointments in order to focus on the overall progression patterns. The information on events and times was set into relation with patient characteristics at the baseline visit by applying different survival analysis techniques, which were applied separately to each SCA type.

#### Evaluation

The reliability of relationships derived from the training data can be checked by predicting the progression risk on unseen data and evaluating the predictions with respect to the observed ground truth. A standard evaluation measure in survival tasks is the concordance index, which is the fraction of correctly ranked comparable pairs among all comparable pairs. A pair of subjects is comparable if either both subjects had a deterioration event and one of them happened earlier than the other, or if one subject had an event and the other subject was observed for at least the same time period without having an event. Subjects with tied events are not comparable. For the concordance index computations in this work we used the function concordance.index from the survcomp R/Bioconductor package, version 1.42.0. A four-fold cross-validation approach was employed for evaluation, splitting the subjects into four groups and leaving each group once as a test set while training the prediction models on the pooled set of the three other groups. Fixed subject groups were used to evaluate different prediction approaches for different neurological symptoms of a specific SCA type. The groups were determined such that each group had a fair share of comparable pairs. For that, a graph-based approach was applied, connecting comparable subjects by edges and recursively splitting the graph into two parts by the graph clustering method cluster\_fast\_greedy from the igraph R package, version 1.2.11. Since the INAS scales yielded a smaller number of comparable pairs than the SARA scales in our data, we took the comparable pairs for the INAS count score to determine the common subject groups in the analyses that included progression of all the scales.

**Table S1.** Statistics for disease progression prediction across all neurological symptoms (37 in total). The given prediction performance is the median concordance index on test data from four-fold cross validation (Figure S5). Progression of a specific symptom is considered to be predicted above random if the concordance index was greater than 0.5 in all test sets of the cross-validation (Figure S6). Progression of a specific symptom is considered dependent on its own baseline if the baseline value is among top three progression predictors of that symptom (Figure S8). For the top progression predictors across all symptoms, we give the percentage of symptoms for which they are among the top three predictors. Low progression INAS items are those among the 26 single INAS items that have less than 5 % progression frequency (Figure S4). Specifically progressing INAS items are those that progress with frequency at least 5 % in only one SCA type.

|  | SCA1 | SCA2 | SCA3 | SCA6 |
| --- | --- | --- | --- | --- |
| Median performance for disease stage progression prediction | 0.76 | 0.76 | 0.78 | 0.80 |
| Percentage of symptoms predicted above random | 58 | 74 | 97 | 55 |
| Percentage of symptoms dependent on own baseline | 59 | 49 | 78 | 51 |
| Top progression predictors across symptoms | SARA sum (46%)<br>stage (35%)<br>CAG expanded (32%) | SARA speech (22%)<br>SARA gait (22%)<br>CAG repeats normal (22%) | SARA sum (41%)<br>SARA gait (35%)<br>CAG expanded (24%) | SARA gait (22%)<br>SARA finger chase (22%)<br>SARA stance (19%) |
| Percentage of low progression INAS items | 23 | 31 | 15 | 54 |
| Specifically progressing INAS items | - | rigidity | dysphagia | downbeat |

#### Progression prediction methods

We applied several different survival prediction methods and comparatively evaluated their performance by the concordance index on the same test sets, considering the time to the first occurring event as described above. First, we computed standard Cox regression using the coxph function from the survival R package, version 3.2-13. This approach trains only on the first occurring event per subject. Second, multiple recurring progression events per subject were taken into account using the Anderson-Gill extension of the Cox model also implemented in the coxph function. Third, a regularized prediction model was obtained by elastic net Cox regression with the the R package glmnet, version 4.1-3, setting family= “cox” and alpha=0.5. Finally, a survival forest approach was performed with the randomForestSRC R package, version 3.0.2, using ntree=1000 and nodesize=10 in the rfsrc function.

#### Feature selection

Additionally, progression events of individual disease stage transitions were modeled for each SCA type. In this analysis, only subjects at one specific disease stage were considered, and a progression event was defined as the transition to a higher disease stage. For evaluation, four-fold cross-validation was applied again, using the comparable pairs with respect to this stage progression for graph-based subject grouping. For each training set, a bootstrapping approach was used to select the most predictive features on hundred resampled instances of the training set. On the one hand, we selected the top five features that most frequently got one of the top five importance scores in a model using all the features. On the other hand, we selected the top five features that most frequently ended up in the feature subset that was determined by a greedy iterative approach adding one feature at a time, namely the one with the best concordance index on a validation set containing 25 % of the current training set. The selected features of each method were subsequently used to train a model on the full training set and test its performance on a left-out test set. In this way, the feature selection approach was performed four times independently on different, partly overlapping training datasets. The features that were selected more than one time were chosen as candidates for further analysis. To understand the roles of these features in the prediction of stage transitions and in particular distinguish positively and negatively influencing factors, a decision tree was learned on all subjects with complete observations for these features. This was done with the R package tree, version 1.0-41, considering a classification into subjects undergoing the stage transition within three years and those that do not. We set mincut=3 and minsize=10 in tree.control. For the mindev parameter, we used the default value of 0.01.

### Supplementary data: Clinical and demographic characteristics

List of neurological scale items and disease stage mapping

**Table S2.** List of items. Comprehensive list of INAS^4^ and SARA^3^ items as well as disease stages^12^ included in the analysis with the used abbreviation and item description.

| Abbreviation | Description |
| --- | --- |
| INAS acuity | Impaired visual acuity (<0.6 for binocular sight in distance testing); INAS item 25 |
| INAS areflexia | Areflexia in either biceps, patellar or achilles tendon reflex; INAS items 1-3 |
| INAS atrophy | Muscle atrophy in either face/tongue, proximal/distal upper/lower limbs; INAS item 7 |
| INAS brainstem ocul. | Ophthalmoparesis on horiz.or vertical gaze and/or slowing of saccades; INAS items 20-21 |
| INAS chorea/dyskinesia | Chorea/Dykinisia in either face/tongue, neck, trunk, upper/lower limbs; INAS item 11 |
| INAS cognitive | Cognitive impairment, according to examiner; INAS item 29 |
| INAS diplopia | Double vision; INAS item 26 |
| INAS downbeat | Downbeat-nystagmus on fixation; INAS item 17 |
| INAS dysphagia | Dysphagia; INAS item 27 |
| INAS dystonia | Dystonia in either face/tongue, neck, trunk, upper/lower limbs; INAS item 12 |
| INAS extensor plantar refl. | Extensor plantar reflex; INAS item 4 |
| INAS fasciculations | Fasciculations in either face/tongue, upper or lower limbs; INAS item 8 |
| INAS gaze nyst. horizontal | Gaze evoked-nystagmus on horizontal testing; INAS item 18 |
| INAS gaze nyst. vertical | Gaze evoked-nystagmus on vertical testing; INAS item 19 |
| INAS hyperreflexia | Hyperreflexia in either biceps, patellar or achilles tendon reflex; INAS items 1-3 |
| INAS hypermetric saccades | Hypermetric saccades; INAS item 24 |
| INAS hypometric saccades | Hypometric saccades; INAS item 23 |
| INAS square wave jerks | INAS square wave jerks on fixation; INAS item 16 |
| INAS myoclonus | Myoclonus in either face/tongue, trunk, upper or lower limbs; INAS item 9 |
| INAS paresis | Paresis in either face/tongue, proximal/distal upper/lower limbs; INAS item 6 |
| INAS pursuit | Broken up smooth pursuit; INAS item 15 |
| INAS rigidity | Rigidity either axial, upper or lower limbs; INAS item 10 |
| INAS sensory | Impaired vibration sense malleolus left or right; INAS item 14 |
| INAS spasticity | Spasticity either in gait, upper or lower limb ; INAS item 5 |
| INAS tremor | Resting tremor; INAS item 13 |
| INAS urinary | Urinary dysfunction; INAS item 28 |
| INAS count |  |
| SARA gait | SARA item 1) Gait |
| SARA stance | SARA item 2) Stance |
| SARA sitting | SARA item 3) Sitting |
| SARA speech | SARA item 4) Speech |
| SARA finger chase | SARA item 5) Finger chase |
| SARA nose-finger | SARA item 6) Nose-finger test |
| SARA fastalhand | SARA item 7) Fast alternating hand movements |
| SARA heel-shin | SARA item 8) Heel-shin slide |
| SARA sum score |  |
| Disease stage | normal, ataxic/gait disturbances, walking aid, wheelchair (doi:10.1093/brain/121.4.589) |
| CAG expanded /normal | number of CAG repeats in the expanded/normal allele |

**Table S3.** Disease stage mapping between the different cohorts. Throughout this work, we use the disease stage annotation from ESMI, EUROSCA and SCA-Registry (first column).

| Disease stage | Corresponding CRC disease stages |
| --- | --- |
| 0: Normal | 0.0-1.0 |
| 1: Gait disturbances | 1.5-2.5 |
| 2: Requiring walking aids | 3.0-4.5 |
| 3: Requiring a wheelchair | 5.0-6.0 |

### Supplementary data: SCA population shows a continuous spectrum of disease severity correlated to the SARA sum score

**Figure S1.**
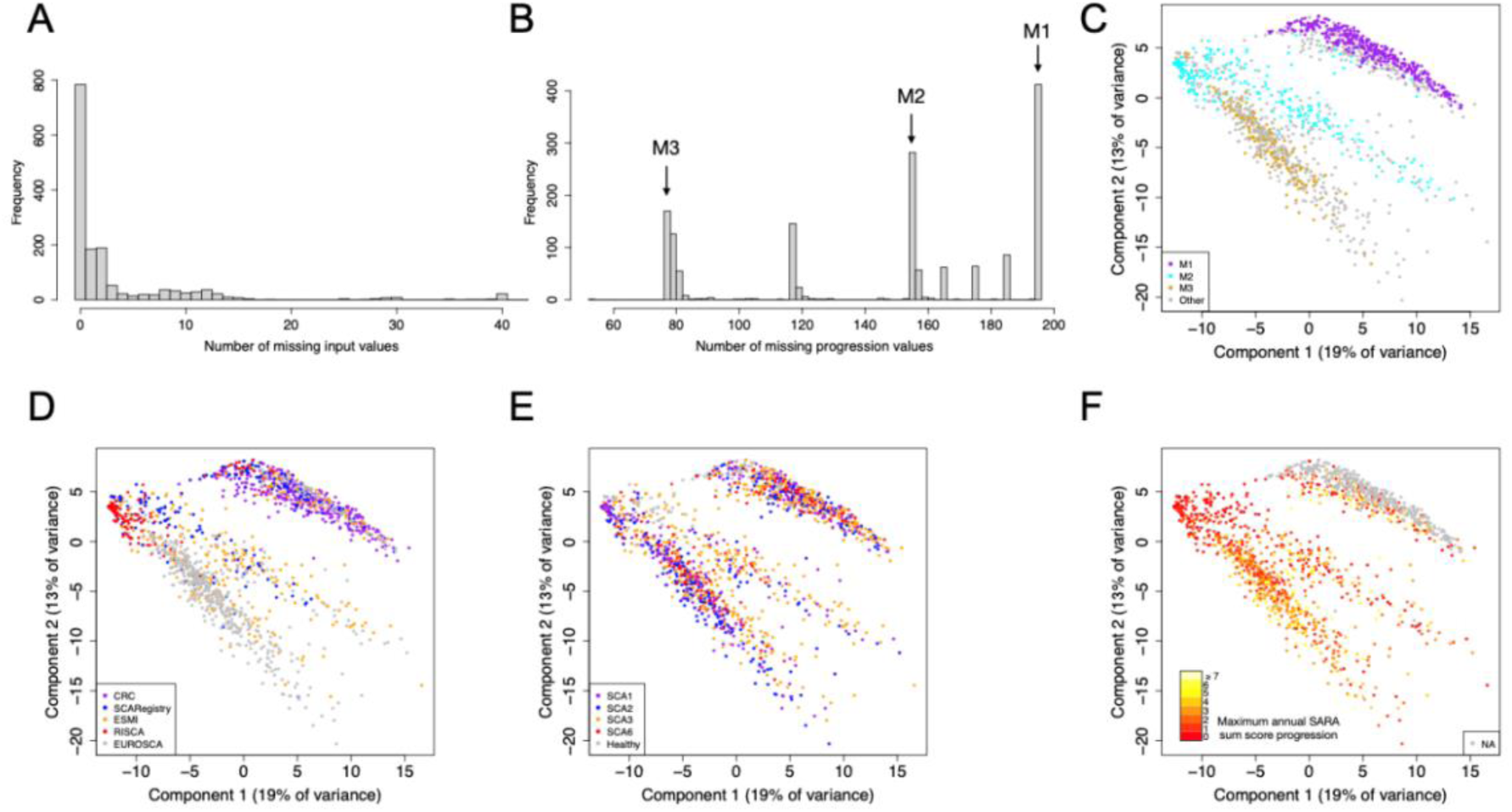
Missing value characteristics of subjects. A. Distribution of numbers of missing values for the input data. The x-axis denotes the number of missing values, the y-axis denotes the number of subjects for each x-axis bin. B. Distribution of numbers of missing values for the progression data (measurements from follow-up visits). The three largest bins are labeled M1, M2 and M3. C. Visualization of subjects based on principal component analysis of clinical time courses across all neurological scales (same as Fig. 1C). Subjects are colored by the major missing value bins (Figure B). D. Visualization of subjects colored by cohort. All CRC subjects belong to the cluster dominated by M3. E. Visualization of subjects colored by disease type. F. Visualization colored by the maximum annual progression rate of SARA sum score. The distribution of missing value clusters is mainly reflected by different cohorts with higher number of missing values. But, there is no relationship between the disease type or annual SARA progression and the missing value structure, disease types and SARA annual progression rates are distributed across all missing value clusters.

**Figure S2.**
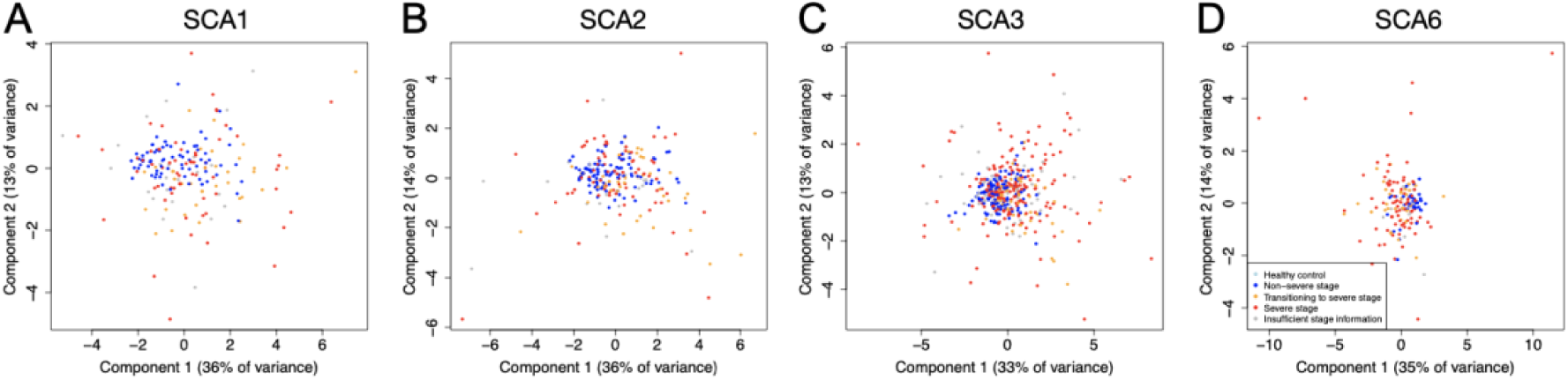
Principal component visualization of subjects based on maximum annual progression rates of the SARA sum score and individual SARA items for each SCA type. Subjects are colored by disease stage annotation. A. Annual progression rate principal component visualization of SCA1 subjects only. After restricting the data to SCA1 subjects. The coloring scheme is the same as before (Figure B). B. Annual progression rate principal component visualization of SCA2 subjects only. C. Annual progression rate principal component visualization of SCA3 subjects only. D. Annual progression rate principal component visualization of SCA6 subjects only.

**Figure S3.**
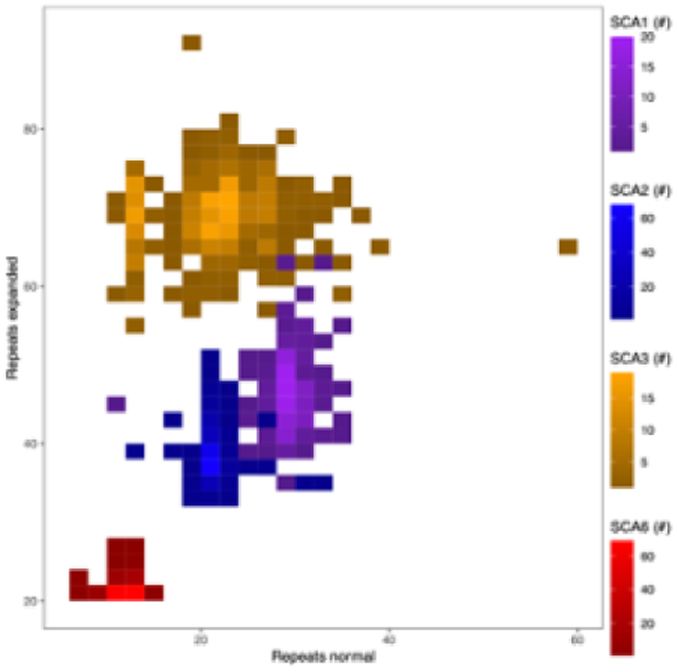
Distribution of numbers of repeats in the normal and the expanded allele. For each bin, the number of subjects with a corresponding repeat number combination is given (\#). The SCA types, marked by different colors, largely occupy different regions in the combination space.

### Supplementary data: Future disease progression events are predictable from the current neurological status

#### Progression frequency

**Figure S4.**
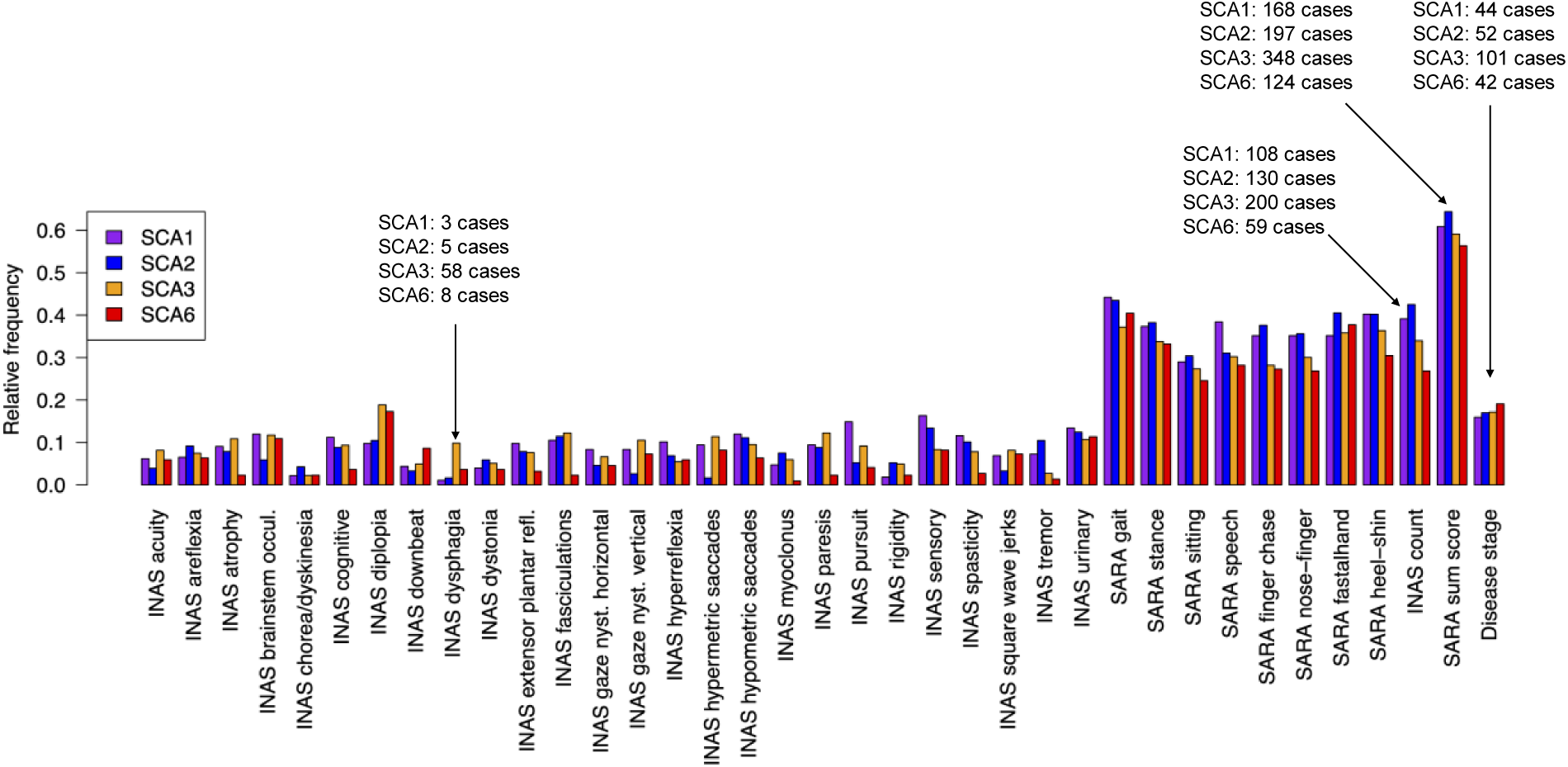
Progression frequency of neurological symptoms among the major SCA types. For each item, the plot indicates the relative fraction of subjects from a specific SCA type showing a deterioration of the respective item within three years from the baseline visit. In addition, the absolute number of cases with progression is indicated for the three summary items and the INAS item with lowest relative progression frequencies in SCA1 and SCA2.

### Supplementary data: Progression predictability

#### Evaluation of methods for predicting disease stage progression

**Figure S5.**
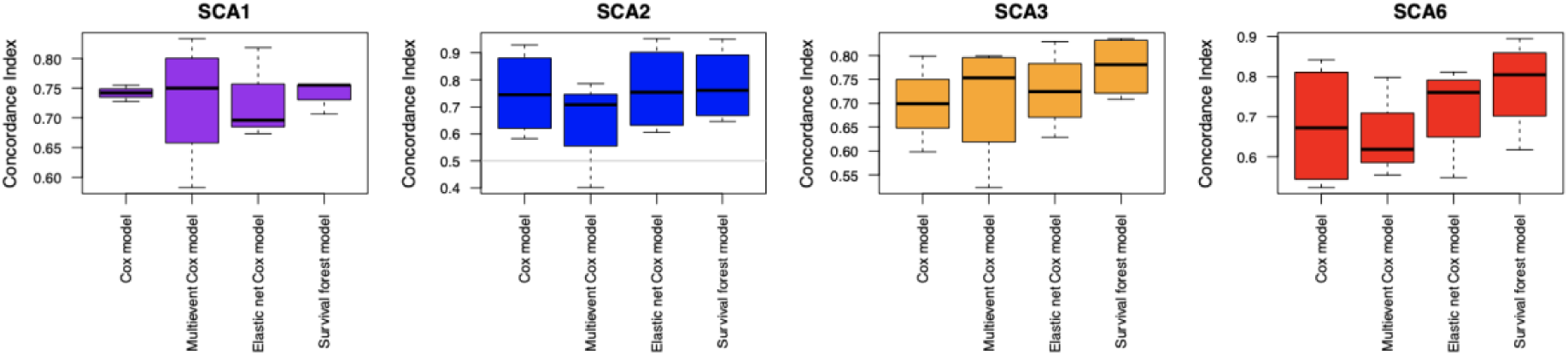
Comparative performance evaluation of methods predicting disease stage progression events individually for each of the most common SCA types. The applied performance measure is the concordance index that assesses the fraction of comparable pairs that were correctly ranked by the prediction approach with respect to the first deterioration event. Random predictions yield a concordance index of 0.5 (marked by the grey line). Boxplots show the distribution of performance values on left-out test data from four-fold cross-validation (bold black line: median, box: 0.25 and 0.75 quantile, whiskers: minimum and maximum value). The following four prediction methods were applied: Cox regression, Cox regression taking all deterioration events of the observed time course into account during training, Cox regression with elastic net regularization to impose usage of a limited number of input features, survival forests. In contrast to Cox regression, survival forests allow for nonlinear combinations of input features. The survival forest approach was the only one that performed consistently well for all the SCA types.

#### Progression predictability for all neurological items and disease stage

**Figure S6.**
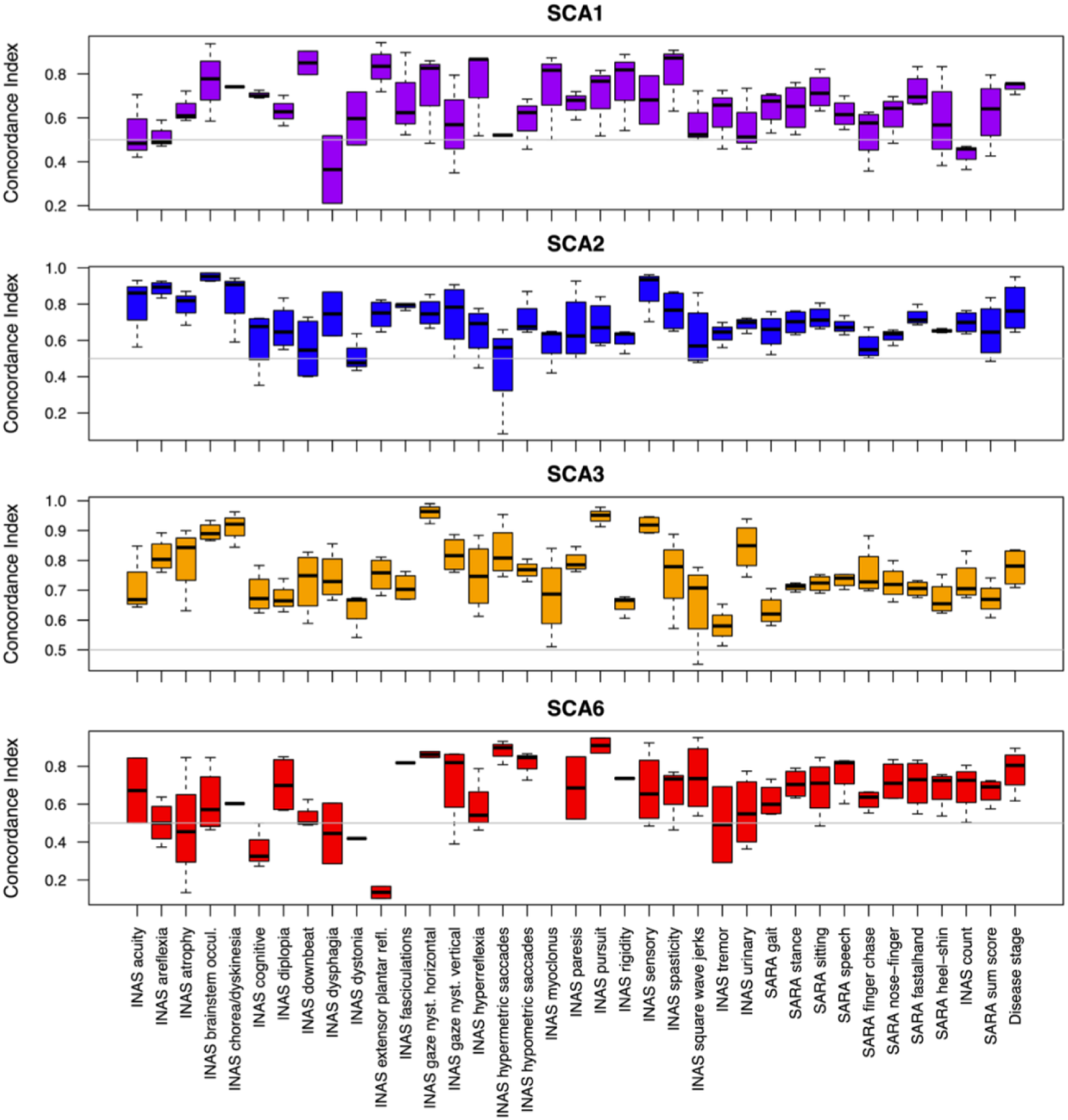
Progression predictability of all neurological items as well as disease stage in each SCA type. Boxplots show the four-fold cross-validation performance of symptom-specific progression prediction by survival forests, indicating how well the progression of each item could be predicted. Random predictions yield a concordance index of 0.5 (marked by the grey line). Progression of all neurological items can be predicted, performing significantly better than random.

### Supplementary data: Identification of the top predictive features

#### Progression prediction for disease stage

**Figure S7.**
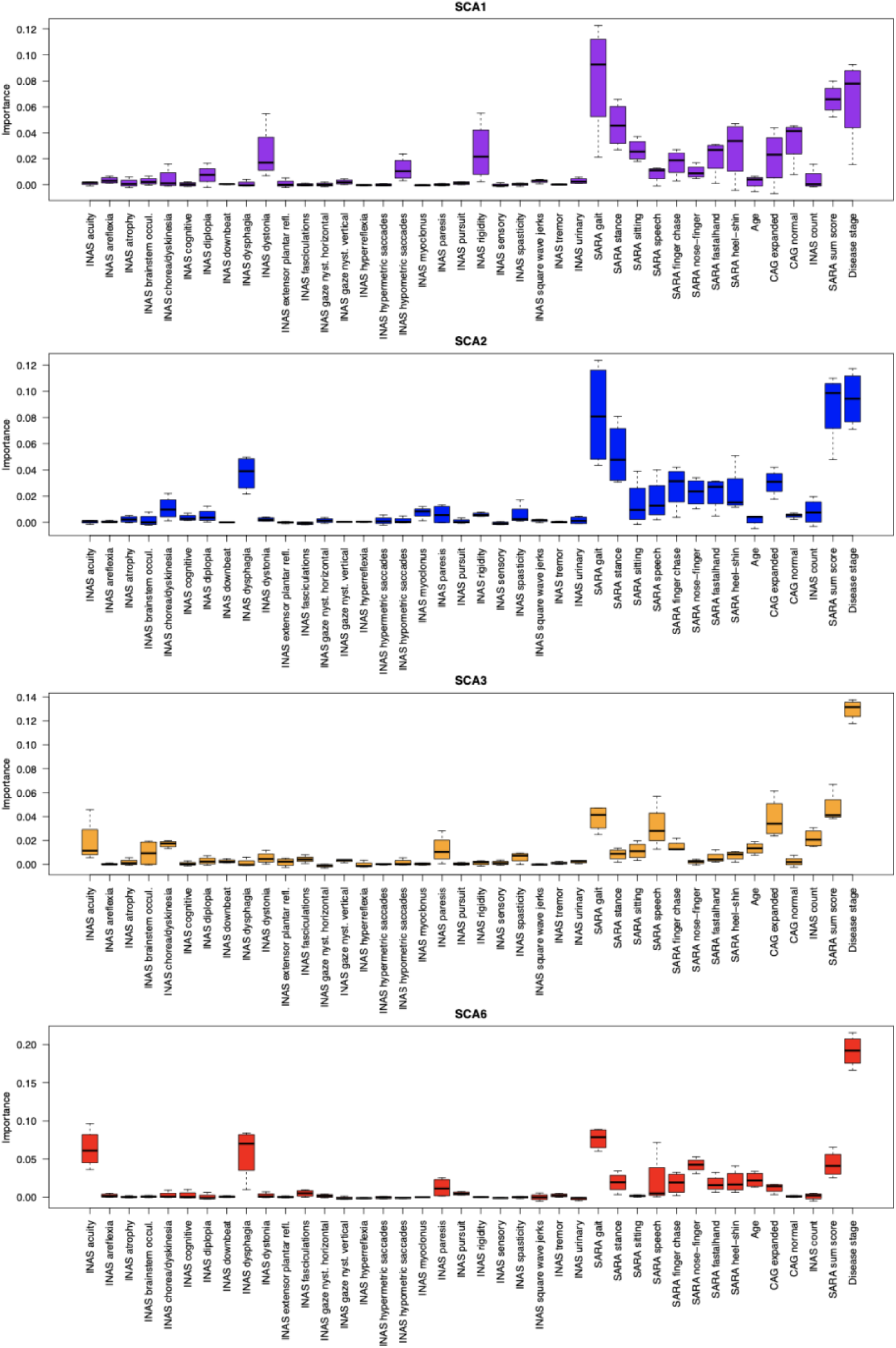
Feature importance for predicting disease stage progression in in SCA1, SCA2, SCA3 and SCA6. The deterioration of disease stage (normal, gait disturbances, walking aids, wheel chair) marks key clinical milestones. For each item the importance values for the prediction of disease stage deterioration were assessed. Boxplots indicate the distribution of importance values assigned by survival forests in four-fold cross-validation for each item (bold black line: median, box: 0.25 and 0.75 quantile, whiskers: minimum and maximum value). A condensed version is given in the main text in Fig. 2}; in which for cross-SCA comparisons of relative feature importances, the top 5 features of each SCA type were selected.

#### Progression prediction of each single item

**Figure S8.**
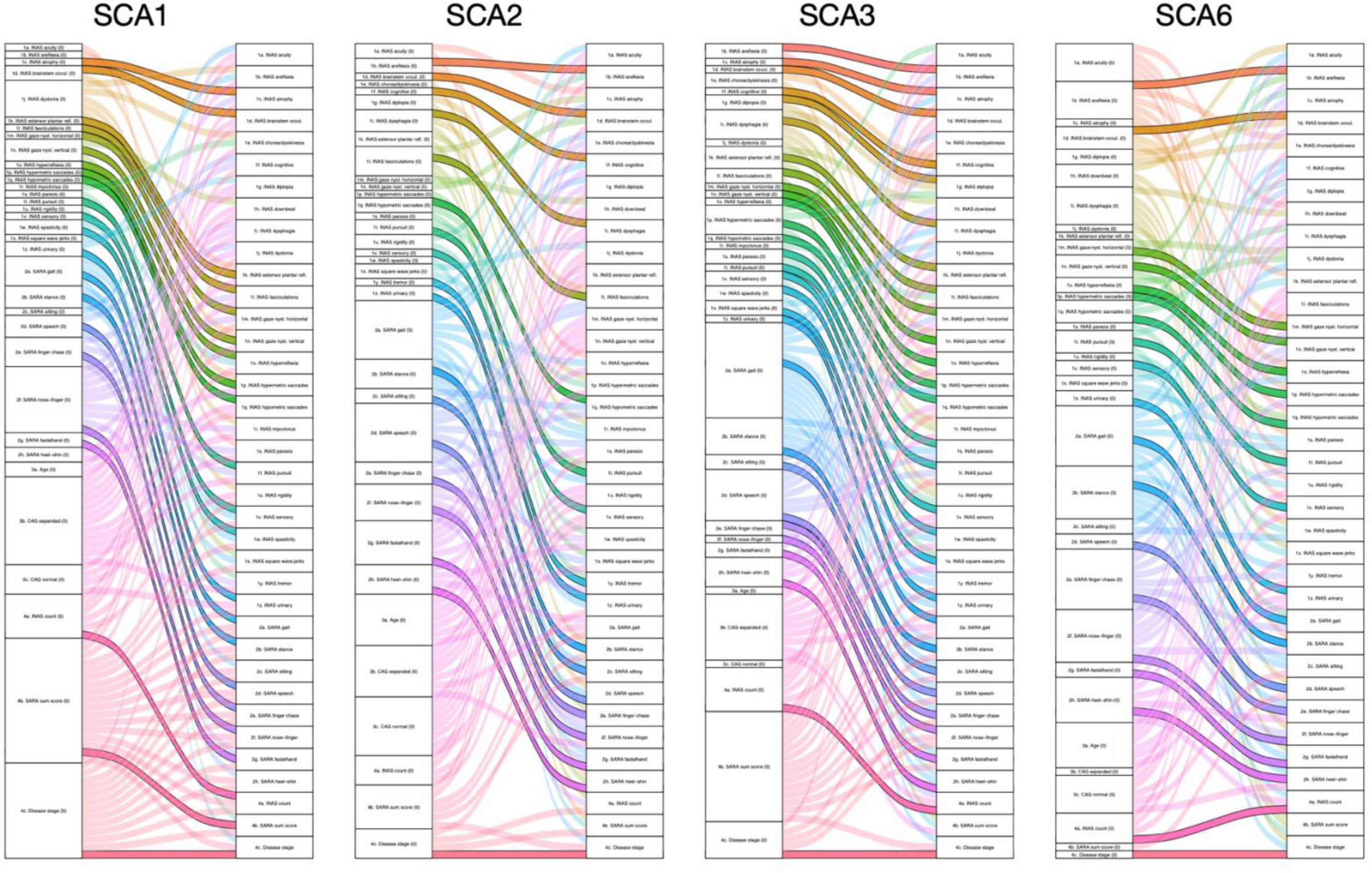
Sankey diagrams representing the top three predictors for the progression of each neurological item in SCA1, SCA2, SCA3 and SCA6.}. The importance values of input features for the prediction of symptom deterioration were assessed in each SCA separately. Neurological measurements at the baseline visit, indicated by the time point zero (0), were the input features for progression prediction. For each item, the features with the top three median importance values were selected. They are represented by the three incoming edges for each progression item in the right column. Edges in stronger color with black borders indicate that progression of a specific neurological scale is predicted by its own baseline value, assessed at the baseline visit. In the left column, the height of each item corresponds to the number of items for which this particular feature was amongst the top three predictors. The majority of scales (74 %) had their own baseline value among the top three predictors (Figure S8, Table S1). Beyond those evident predictors, there were many cross-relationships among items. To outline one particular example, the SARA item gait prediction for SCA3 had a median concordance index of 0.62 and its top three predictors were its own baseline value, SARA sum and CAG repeats of the expanded allele. While this example might be rather expected, in other cases, combinations emerge, that are less to be anticipated. Interestingly, the top three predictors for the SARA sum score in SCA3 and SCA6 were non-SARA items. Overall, the baseline score of SARA gait was among the most universal predictors across symptoms for SCA2, SCA3 and SCA6 (Table S1). Likewise, the number of repeats in the expanded allele was a main predictor for SCA1 and SCA3. Top progression predictors across symptoms.

**Figure S9.**
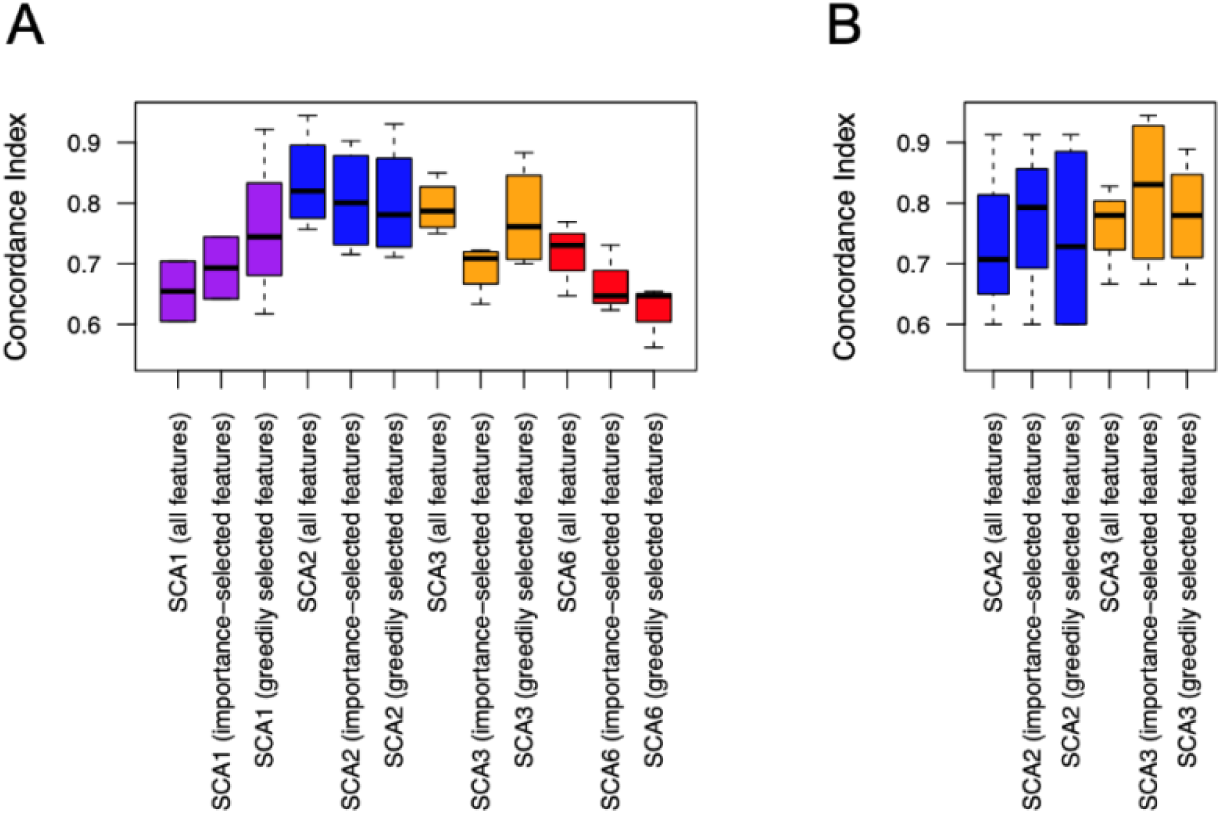
Cross-validation performance of survival forest models predicting specific disease stage transitions. A. Prediction performance for the transition from disease stage 1 to disease stage 2 (loss of free walking ability). In addition to the performance of models using all input features, the boxplots show the performance of models using a set of five features that were selected by a bootstrap on the respective training set, based either on importance scores or on a greedy iterative procedure (Methods). B. Prediction performance for the transition from disease stage 2 to disease stage 3 (loss of ambulation). Due to the low number of cases (less than twelve transitions), cross-validation performance is not reported for SCA1 and SCA6.

### Supplementary Data: Comparison of SCA types and cross-validation of models

#### Cross-Validation of models

Transition from disease stage 1 to 2 and 2 to 3 in SCA3 are shown in Figure 3 and 4, respectively, and we now compare them to the respective stage transitions of the other SCA types with less observational points available. While SCA2 and SCA3 maintained high performance levels similar to the overall stage progression prediction, SCA1 and SCA6 with their lower number of cases showed a performance drop when specifically predicting the loss of free walking transition (Figure S9 A). For SCA1, the prediction performance was rescued when restricting the model to the top five predictive features greedily selected on the training data. In contrast, SCA3 kept a similar performance and SCA2 and SCA6 showed a worse performance for the restricted models. Regarding the risk of needing a wheelchair however, predictions for SCA2 and SCA3 seemed to benefit from restricting the models to five predictive features, in particular when selecting the features based on their importance in the large model with all features (Figure S9 B). We conclude that generally, independent of the specific stage transition of interest, it is instructive to consider the whole feature profiles during predictions. Only in the case of very small training datasets, such large models may overfit and feature selection may improve the prediction performance.

For SCA1 and SCA6, there were less than twelve patients undergoing the transition from stage 2 to stage 3. Therefore, it was not possible to test the model generalizability by cross-validation.

Nevertheless, we performed for each SCA type and stage selection the same training set selection and feature selection procedures (Methods) and compared the resulting stage-1-to-2 models (Figures S10, S11, S13 with the stage-2-to-3 models (Figures S14, S15, S17) and the cross-stage models (Figure 2).

Please note, for SCA3 charts are given with the additional information of cumulative hazard and respective frequencies (Figures S12, S16) that were not included in the figures in the main manuscript (Figures 3, 4).

#### Specific disease stage transitions

**Table S4.**
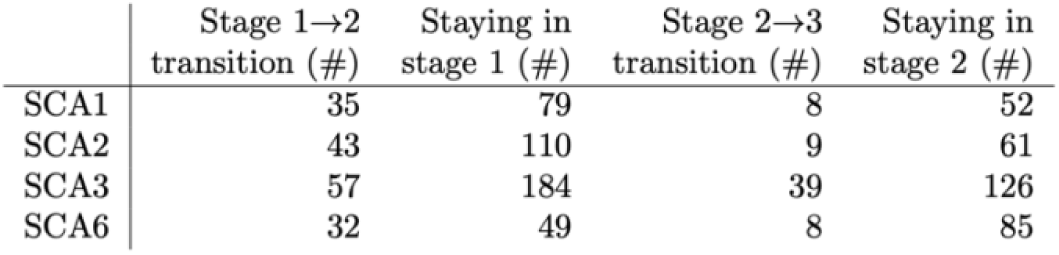
Summary of stage transitions. Number of subjects of each disease type for which follow-up visits existed and a corresponding transition was observed or not.

|  | Stage 1→2<br>transition (#) | Staying in<br>stage 1 (#) | Stage 2→3<br>transition (#) | Staying in<br>stage 2 (#) |
| --- | --- | --- | --- | --- |
| SCA1 | 35 | 79 | 8 | 52 |
| SCA2 | 43 | 110 | 9 | 61 |
| SCA3 | 57 | 184 | 39 | 126 |
| SCA6 | 32 | 49 | 8 | 85 |

The decision trees are solely shown for ease of interpretation and do not constitute the full prediction models.

#### Background and interpretation of figures

Each figures aims to assess the total risk for patients of transitioning within three years from disease stage 1 to disease stage 2 (loss of free walking ability) or disease stage 2 to 3 (loss of ambulation), respectively.

The risk was quantified in the context of robustly identified top predictive features. In other words, each column on the left hand side represents one of the features, that were robustly identified as a main contributor to the prediction for the loss of free walking ability within the next 3 years. Color coding of each feature range is indicated by the Color Key under the respective column with blue coloring indicating low values and red coloring indicating high values. The risk for progression in terms of the cumulative hazard was computed based on the whole input feature profiles by the survival forest approach. The top predictive features were selected by training set bootstrapping within a four-fold cross-validation (Methods, Figure S9). The cumulative hazard was divided into bins of width 0.2, and for each bin the number of cases (Histogram next to the cumulative hazard column) and the median value of each predictive feature is shown (feature columns on the left side).

The graph allows interpretations in horizontal as well as subsequently vertical reading. Following a horizontal line, one particular combination of features can be related to the respective predicted 3-year outcome by following a imagined horizontal line to the right-hand side, in particular, to the resulting probability to keep the ability of free walking within three years (given with the column on the right-hand side), or in other words a high or low risk for the loss of free walking ability (indicated by the triangle). E.g. following the 2 bottom bins, one can see that low values in each features are related to a low risk and almost 100 % probability to keep the ability of free walking within the next 3 years. A vertical reading of the columns allows to identify patterns within single features related to the risk of losing the ability of free walking. It becomes obvious that, e.g. in general higher values in the SARA sum score lead to a high risk of deterioration. However, this hold true for other disease specific items, e.g. is the presence of fasciculations in SCA3 associated with a relatively high risk for the loss of the ability to walk freely. Here, higher values always occur in the upper portion of the respective column, which is related to a high risk (as indicated by the probability column as well as the triangle on the right-hand side). In contrast, e.g. the values of age or INAS count are often mixed across the vertical of their columns, with no consistent discernible pattern. Even though, they were identified as the top predictive features, the overall risk of progression depends substantially on the constellation of the other features.

B. To facilitate interpretation decision tress were built using only features that were selected multiple times in different training data sets. The resulting decision trees built from the robustly identified top predictive features illustrate their combined effects on the probability of non-progression within the next 3 years, derived from the mean cumulative hazard in each leaf. They are solely shown for ease of interpretation and do not constitute the full prediction model. At each inner node of each tree, the left branch satisfies the condition written at the top of the node, whereas the right branch does not satisfy it. The probability to remain in disease stage 1 or 2, or in other words keeping the ability for free walking or ambulantion is given at each branching end and is color coded according to the probability on the right-hand side in (A).

### Transition from disease stage 1 to disease stage 2 for SCA1, SCA2, SCA3 and SCA6 patients: Loss of free walking

**Figure S10.**
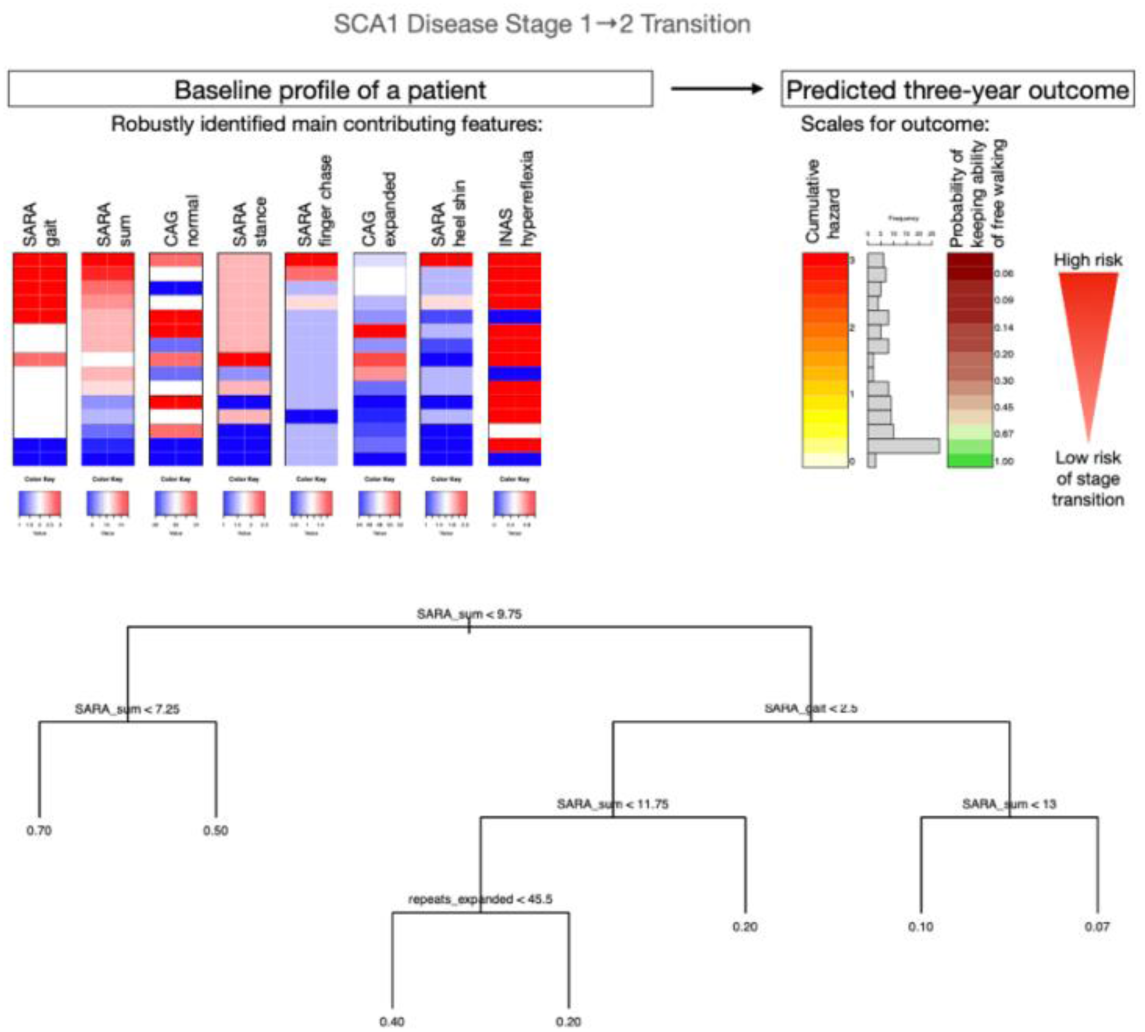
Assessing the total risk for SCA1 patients of transitioning within three years from disease stage 1 to disease stage 2 (loss of free walking ability) in the context of top predictive features. The risk in terms of the cumulative hazard was computed based on the whole input feature profiles by the survival forest approach. The top predictive features were robustly identified by training set bootstrapping in multiple cross-validation folds (see Feature selection subsection in Supplementary Methods). Here, the features are sorted by their importance in a model using all data and features. The cumulative hazard was divided into bins of width 0.2, and for each bin the number of cases and the median value of each predictive feature is shown. The probability of remaining in disease stage 1 is exponentially related to the cumulative hazard. The decision tree was built from the robustly identified top predictive features to illustrate their combined effects on the probability of remaining in disease stage 1, derived from the mean cumulative hazard in each leaf. It is solely shown for ease of interpretation and does not constitute the full prediction model. At each inner node of the tree, the left branch satisfies the condition written at the top of the node, whereas the right branch does not satisfy it.

**Figure S11.**
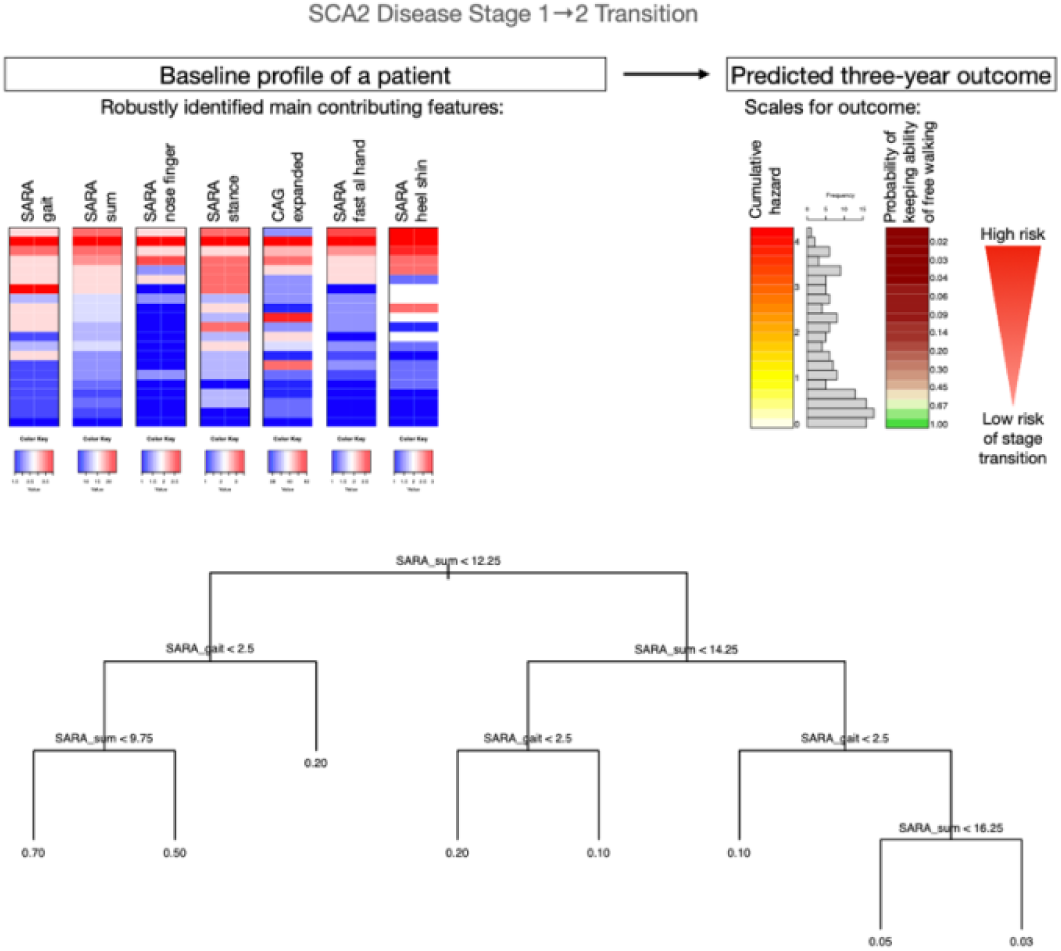
Assessing the total risk for SCA2 patients of transitioning within three years from disease stage 1 to disease stage 2 (loss of free walking ability) in the context of robustly identified top predictive features.

**Figure S12:**
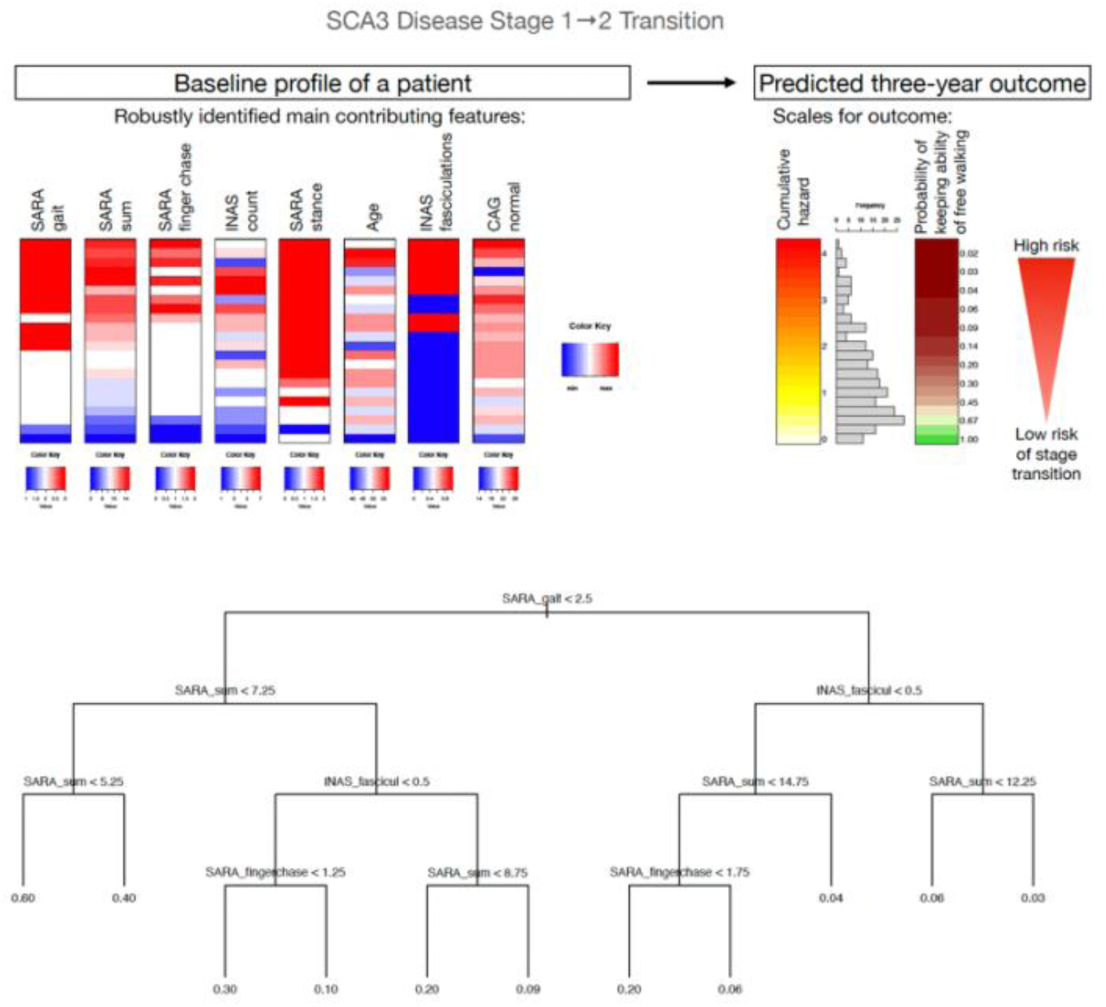
Assessing the total risk for SCA3 patients of transitioning within three years from disease stage 1 to disease stage 2 (loss of free walking ability) in the context of robustly identified top predictive features.

**Figure S13.**
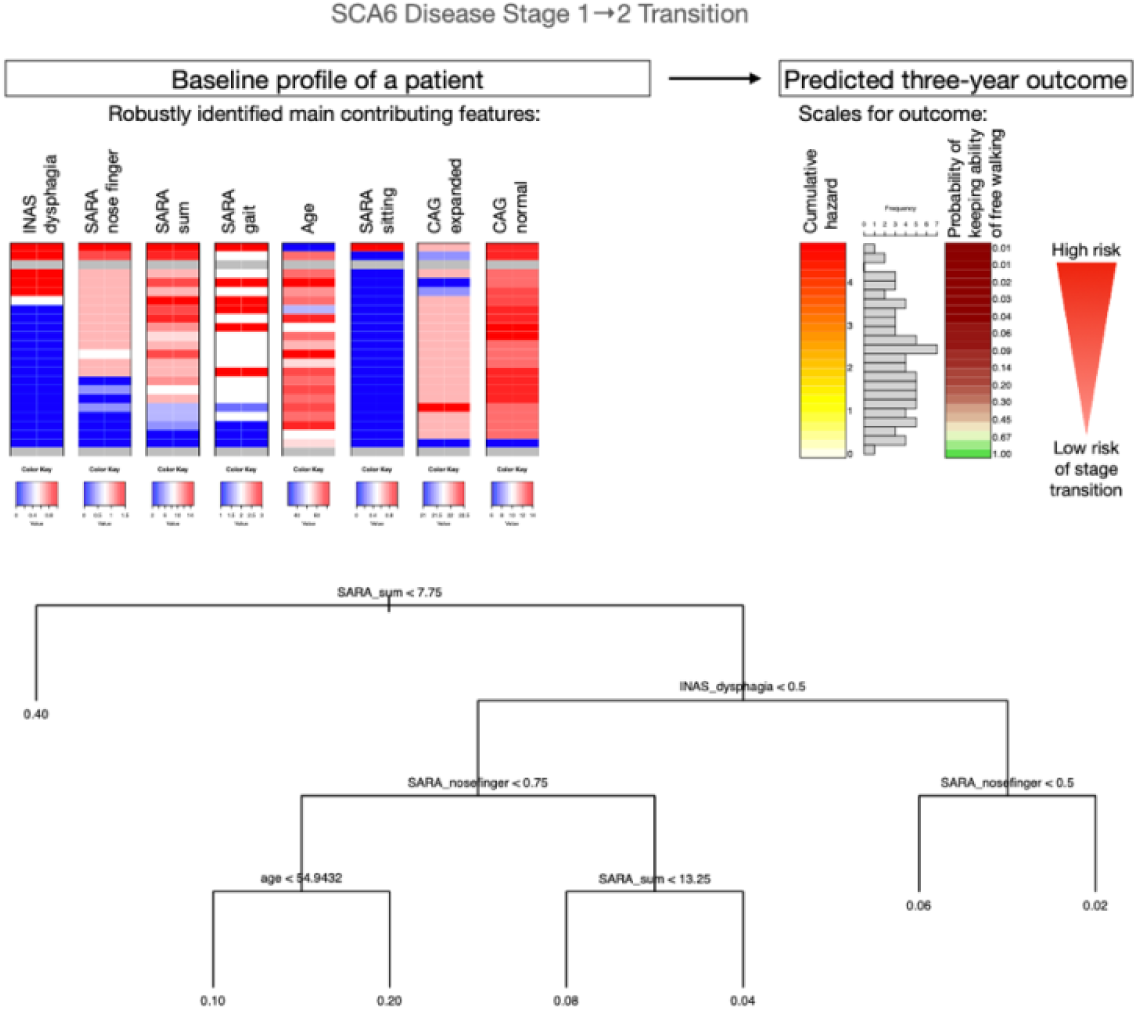
Assessing the total risk for SCA6 patients of transitioning within three years from disease stage 1 to disease stage 2 (loss of free walking ability) in the context of robustly identified top predictive features.

### Transition from disease stage 2 to disease stage 3 in SCA1, SCA2 and SCA6: Loss of ambulation

**Figure S14.**
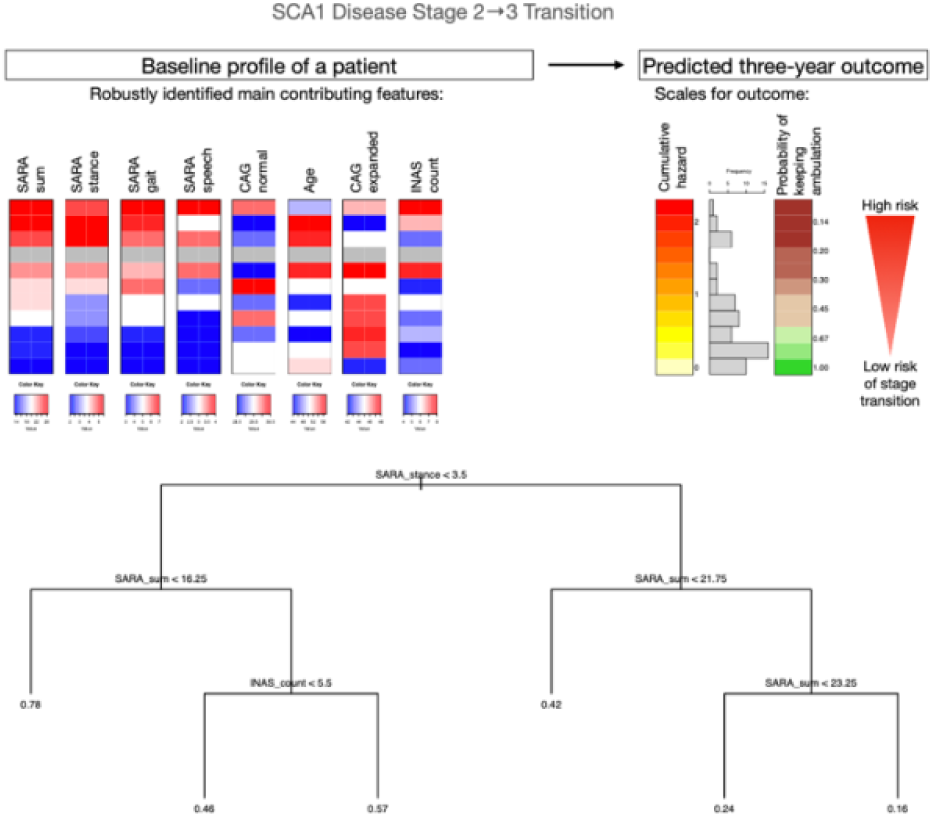
Assessing the total risk for SCA1 patients of transitioning within three years from disease stage 2 to disease stage 3 (need of using a wheelchair) in the context of robustly identified top predictive features.

**Figure S15.**
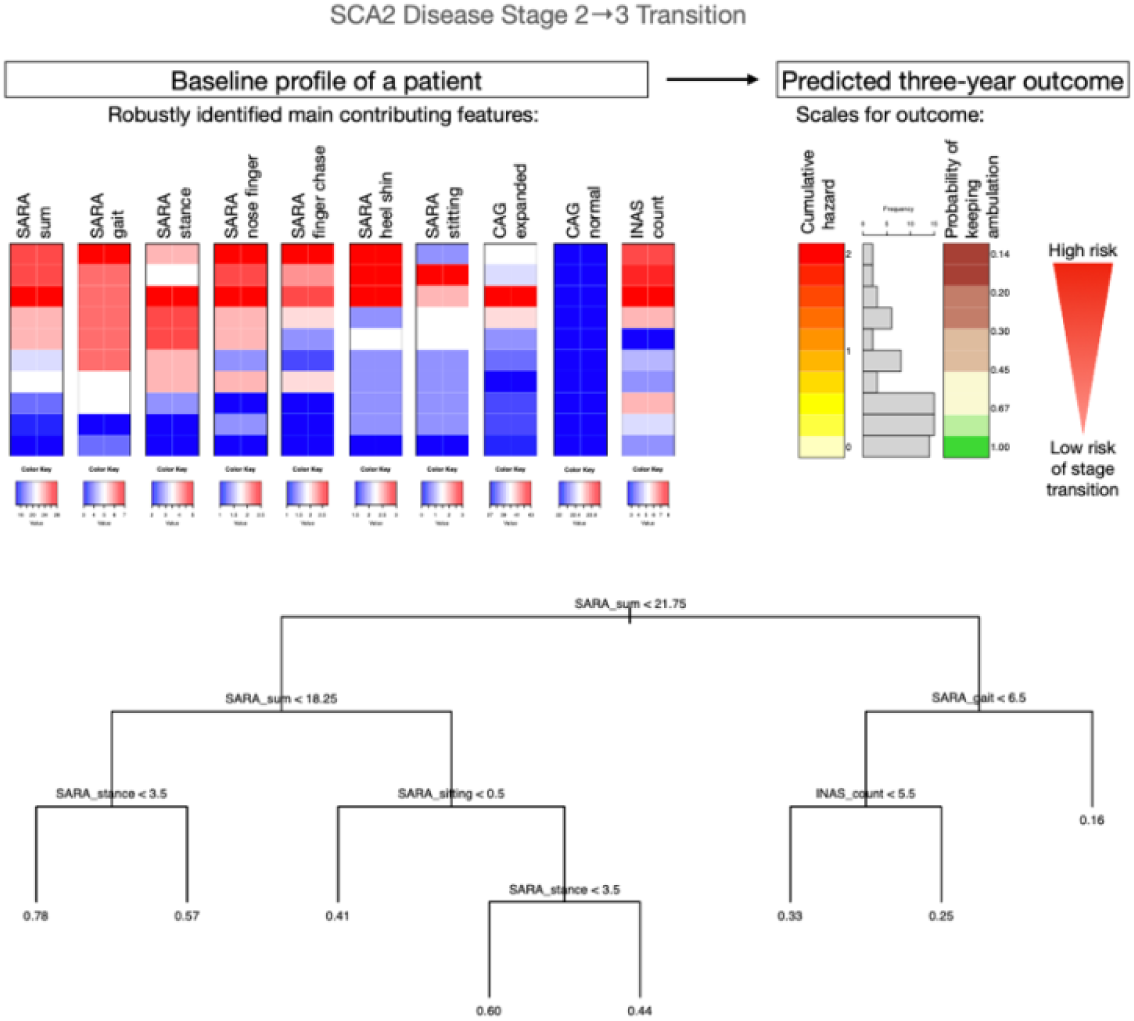
Assessing the total risk for SCA2 patients of transitioning within three years from disease stage 2 to disease stage 3 (need of using a wheelchair) in the context of robustly identified top predictive features.

**Figure S16:**
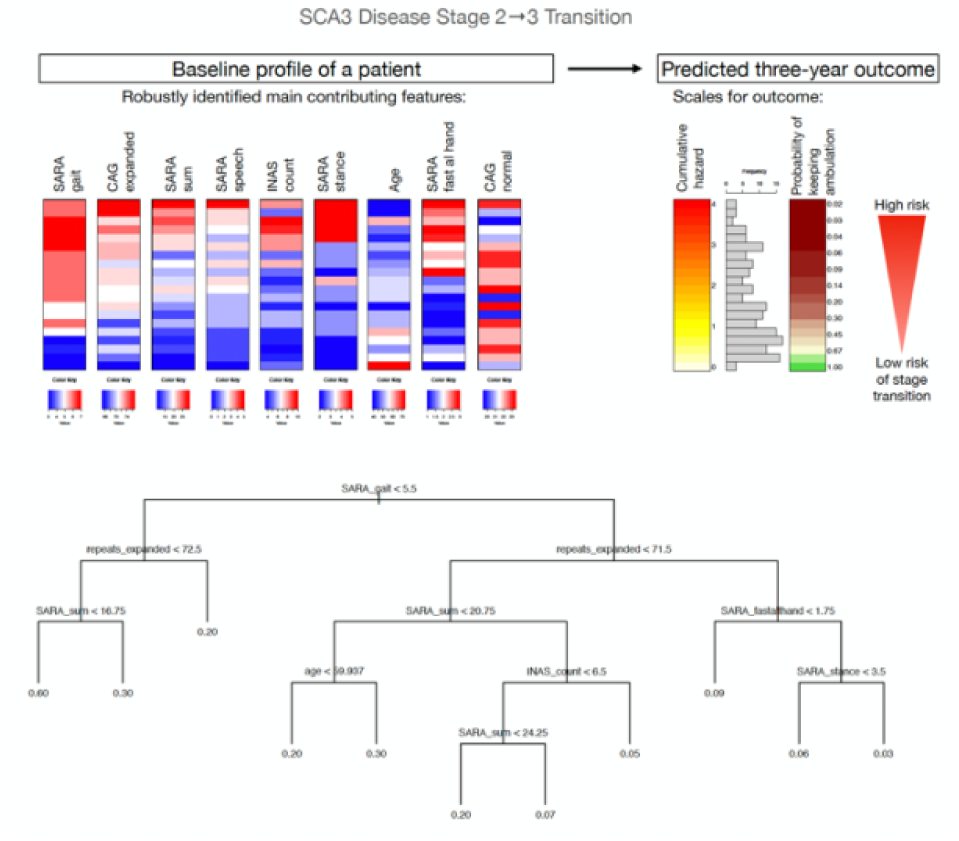
Assessing the total risk for SCA3 patients of transitioning within three years from disease stage 2 to disease stage 3 (need of using a wheelchair) in the context of robustly identified top predictive features.

**Figure S17.**
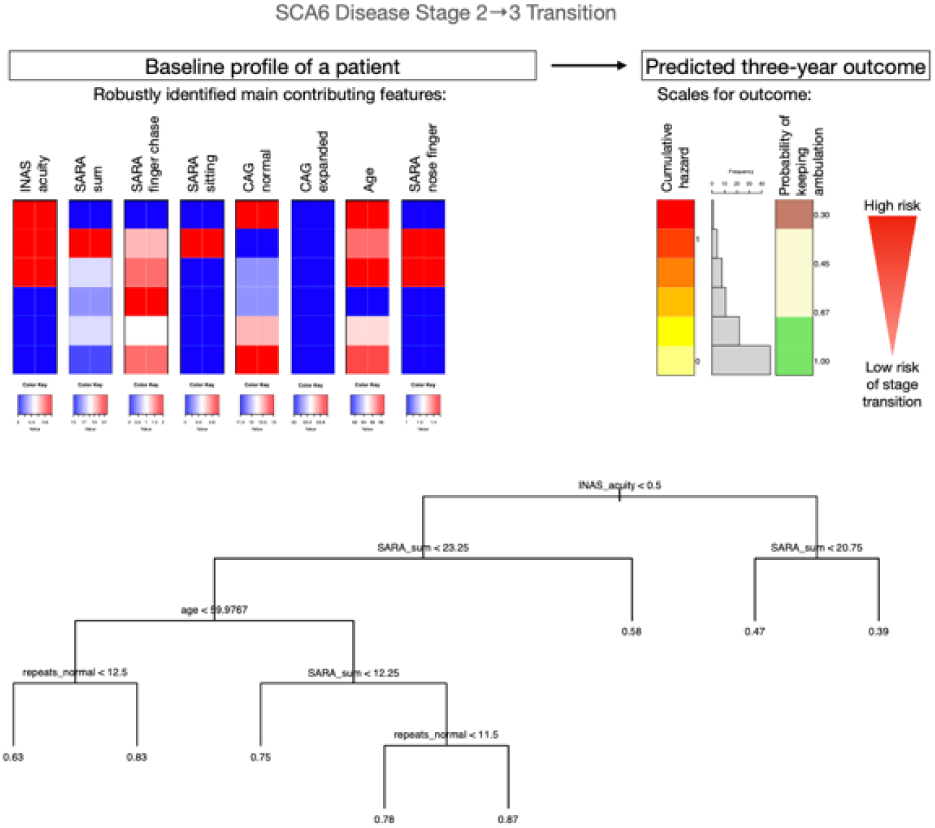
Assessing the total risk for SCA6 patients of transitioning within three years from disease stage 2 to disease stage 3 (need of using a wheelchair) in the context of robustly identified top predictive features.

## Notes

### Competing Interest Statement

The authors have declared no competing interest.

### Funding Statement

This work was funded by the National Ataxia Foundation (NAF). JF received funding by the iBehave Network, sponsored by the Ministry of Culture and Science of the State of North Rhine-Westphalia and as a Fellow of the Hertie Network of Excellence in Clinical Neuroscience.

### Author Declarations

Ethics committee/IRB of the University Hospital Bonn gave ethical approval for this work.

